# Initial data analysis for longitudinal studies to build a solid foundation for reproducible analysis

**DOI:** 10.1101/2023.12.05.23299518

**Authors:** Lara Lusa, Ćecile Proust-Lima, Carsten O. Schmidt, Katherine J. Lee, Saskia le Cessie, Mark Baillie, Frank Lawrence, Marianne Huebner, TG3 of the STRATOS Initiative

**Affiliations:** Department of Mathematics, Faculty of Mathematics, Natural Sciences and Information Technologies, University of Primorska, Koper/Capodistria, Slovenia; Institute for Biostatistics and Medical Informatics, Faculty of Medicine, University of Ljubljana, Ljubljana, Slovenia; Univ. Bordeaux, Inserm, Bordeaux Population Health Research Center, UMR1219, F-33000 Bordeaux, France; Institute for Community Medicine, SHIP-KEF University Medicine of Greifswald, Greifswald, Germany; Clinical Epidemiology and Biostatistics Unit, Murdoch Children’s Research Institute, Melbourne, Australia; University of Melbourne, Melbourne, Australia; Department of Clinical Epidemiology and Department of Biomedical Data Sciences, Leiden University Medical Center, Leiden, The Netherlands; Department of Biomedical Data Sciences, Leiden University Medical Center, Leiden, The Netherlands; Novartis, Basel, Switzerland; Center for Statistical Training and Consulting, Michigan State University, East Lansing, MI, USA; Department of Statistics and Probability, Michigan State University, East Lansing, MI, USA

## Abstract

Initial data analysis (IDA) is the part of the data pipeline that takes place between the end of data retrieval and the beginning of data analysis that addresses the research question. Systematic IDA and clear reporting of the IDA findings is an important step towards reproducible research. A general framework of IDA for observational studies includes data cleaning, data screening, and possible updates of pre-planned statistical analyses. Longitudinal studies, where participants are observed repeatedly over time, pose additional challenges, as they have special features that should be taken into account in the IDA steps before addressing the research question. We propose a systematic approach in longitudinal studies to examine data properties prior to conducting planned statistical analyses.

In this paper we focus on the data screening element of IDA, assuming that the research aims are accompanied by an analysis plan, meta-data are well documented, and data cleaning has already been performed. IDA screening domains are participation profiles over time, missing data, and univariate and multivariate descriptions, and longitudinal aspects. Executing the IDA plan will result in an IDA report to inform data analysts about data properties and possible implications for the analysis plan that are other elements of the IDA framework.

Our framework is illustrated focusing on hand grip strength outcome data from a data collection across several waves in a complex survey. We provide reproducible R code on a public repository, presenting a detailed data screening plan for the investigation of the average rate of age-associated decline of grip strength.

With our checklist and reproducible R code we provide data analysts a framework to work with longitudinal data in an informed way, enhancing the reproducibility and validity of their work.

## 1 Introduction

Initial data analysis (IDA) is the part of the data pipeline that commonly takes place between the end of data retrieval and the beginning of data analysis that addresses the research question. The main aim of IDA is to provide reliable knowledge about the data to ensure transparency and integrity of preconditions to conduct appropriate statistical analyses and correct interpretation of the results to answer pre-defined research questions. A general framework of IDA for observational studies includes the following six steps: (1) metadata setup (to summarize the background information about data), (2) data cleaning (to identify and correct technical errors), (3) data screening (to examine data properties), (4) initial data reporting (to document findings from the previous steps), (5) refining and updating the research analysis plan, and (6) documenting and reporting IDA in research papers [1]. Statistical practitioners often do not perform such necessary steps in a systematic way. They may combine data screening steps with analyses steps leading to ad-hoc decisions; however, maintaining a structured workflow is a fundamental step towards reproducible research [2].

The value of an effective IDA strategy for data analysts lies in ensuring that data are of sufficient quality, that model assumptions made in the analysis strategy are satisfied and are adequately documented, and in supporting decisions for the statistical analyses [3]. IDA data screening investigations could lead to discovery of data properties that may identify further errors in the data, affect the interpretation of results of statistical models, and/or modify the choices linked to the specification of the model.

An IDA checklist for data screening in the context of regression models for continuous, count or binary outcomes was proposed recently [4], not considering outcomes that were of survival-type, multivariate or longitudinal. The goal of this study is to extend the checklist to longitudinal studies, where participants are measured repeatedly over time and the main research question is addressed using a regression model; the focus is on data screening (IDA step 3), where the examination of the data properties provide the data analyst with important information related to the intended analysis. While an investigation of missing data, and univariate and multivariate description of variables is common across studies [4], longitudinal studies pose additional challenges for IDA. Different time metrics, the description of how much much data was collected through the study (how many observations and at which times), missing values across time points, including drop-out, and longitudinal trends of variables should be considered. Model building and inference for longitudinal studies have received much attention [5] and many textbooks on longitudinal studies discuss data exploration and the specific challenges due to missing values [5–7]; however, a systematic process for data screening is missing.

We propose a comprehensive checklist for the data screening step of the IDA framework, which includes data summaries to help understanding data properties, and their potential impact on the analyses and interpretation of results. The checklist can be used for observational longitudinal studies, which include panel studies, cohort studies, or retrospective studies. Potential applications include medical studies designed to follow-up patients in time, electronic health records with longitudinal observations, complex surveys. Other aspects of the IDA framework such as data preparation and data cleaning have been discussed elsewhere ([8], [9]).

This paper is an effort to bring attention to a systematic approach of initial data analysis for longitudinal studies that could affect the analysis plan, presentation, or interpretation of modeling results. This contributes to the general aim of the international initiative STRATOS to strengthen the analytic thinking in the design and analysis of observational studies (http://stratos-initiative.org) [10].

We outline the setting and scope of our paper in Section 2. We describe the necessary steps for data screening of longitudinal studies in section 3, where a check list is also provided. A case study is presented in section 4, using hand grip strength from a data collection across several waves in a complex survey [11]; we present several data summarizations and visualizations, and provide a reproducible R vignette for this application. Possible consequences of the IDA findings for the analyses in this case study are presented in section 4.5, where we discuss the potential implications to the statistical modeling or interpretation of results based on the evaluation of the data properties. The paper ends with the discussion.

## 2 Setting and scope

A plan for data screening should be matched to the research aims, study settings, and analysis strategy. We assume that the study protocol describes a research question that involves longitudinal data, where the outcome variable is measured repeatedly over time, and is analysed using a regression model applied to all time points or measurements. We assume that baseline explanatory variables are measured, and consider also the possibility of time-varying explanatory variables.

“Measurement” in longitudinal studies could refer to a data collection with survey instruments, interviews, physical examinations, or laboratory measurements. Time points at which the measurements are obtained and the number of measurements can vary between individuals. Time series, time-to-event models or applications where the number of explanatory variables is extremely large (omics/high-dimensional) are out-of-scope for this paper. We assume that only one outcome variable is measured repeatedly over time, but most of the considerations would apply also to longitudinal studies with multiple outcomes. We focus on observational longitudinal studies, but most of the explorations that we propose would be appropriate also for experimental studies.

Important prerequisites for a data screening checklist have been described in [4]. A clearly defined research question must be defined, and an analysis strategy for addressing it must be known. The analysis strategy includes the type of statistical model for longitudinal data, defines variables to be considered for the model, expected methods for handling missing data, and model performance measurements. A statistical analysis plan can be built from the analysis strategy and the data screening plan. Structural variables in the context of IDA were introduced in [4]; these are variables that are likely to be critical for describing the sample (e.g. variables that could highlight specific patterns) and that are used to structure IDA results. They can be demographic variables, variables central to the research aim, or levels of measurement (centers); they may or be not also explanatory variables used in the analysis strategy. They help to organize IDA results to provide a clear overview of data properties, in particular limiting the potentially large number of explorations of multivariate distributions, as it is suggested that the association between the structural variables and the explanatory variables is explored [4]. For example, summary statistics stratified by centers might provide valuable information about the data collection process, those stratified by sex or age group might be easier to understand.

We assume that data retrieval, data management and data cleaning (identification of errors and inconsistencies) have already been performed. These aspects comprise specific challenges with longitudinal data, where data sets are prepared in multiple formats (long, one row per measurement, the preferred format for data modeling, and wide, one row per participant, for data visualizations), the harmonization of variable definitions across measurements/over time is often needed, and inconsistencies of repeated measurements across time might be identified in data cleaning. A data dictionary and sufficient meta-data should be available to clarify the meaning and expected values of each variable and information about study protocol and data collection.

An important principle of IDA is, as much as possible, to avoid hypothesis generating activities. Therefore, in the data screening process, associations between the outcome variable and the explanatory variables are not evaluated. However, evaluating the changes of the outcome in time is part of the outcome assessment in the IDA for longitudinal data.

Because longitudinal studies can be very heterogeneous in their data structure, it is challenging to propose a unified data screening checklist. The topics addressed in this paper and summarized in our checklist can be considered a minimum set of analyses to include in an IDA report for transparency and reproducibility to prepare for the statistical modeling to address the research questions; the optional extensions present explorations that might be relevant only in some studies.

## 3 IDA data screening checklist for longitudinal data

To address the specificities of longitudinal studies we extend the IDA data screening checklist proposed for regression modeling [4], which included three domains: missing data, univariate descriptions, multivariate descriptions. In our work the missing values domain is substantially extended, the univariate and multivariate descriptions include explorations at time points after baseline, and two new domains are included: participation profile and longitudinal aspects.

Several items of the IDA screening checklist suggest to summarize data for each time point, which is sensible for study designs where all the individuals have pre-planned common times of measurements or when the number of different times is limited; in this case these times can be used as structural variables in IDA (for instance time visits or waves). For studies where the time points are many and/or uncommon, or not determined by design (random times of observation), we suggest that, for description purposes, the time metric is summarized in intervals and the summaries are provided by time intervals rather than for each of the time points.

The aims of the IDA screening domains and the main aspects of each domain are presented in the following sections, and summarized in Table 1.

**Table 1.** Initial Data Analysis checklist for data screening in longitudinal studies.

| Topic | Item | Features |
| --- | --- | --- |
| **IDA screening domain: Participation profile** |  |  |
| Time frame | P1 | Provide number of time points and intervals at which measurements are taken, using the time metric that best reflects the time from inclusion in the study, or calendar time in studies that involve long enrollment times. Highlight the differences between the time of first measurements and follow-up times. |
| Time metric | P2 | Describe the time metric and corresponding time points specified in the analysis strategy, if different from the time metric described in P1. |
| Participants | P3 | Provide the number of participants who attended the assessment by time metric(s). |
| Optional extensions: Participation Profile |  |  |
| Other time metrics | PE1 | Use different time metric(s) to describe the time frame of the study, if applicable and appropriate, e.g. calendar time or data collection visits. |
| **IDA screening domain: Missing data** |  |  |
| Non-enrollment | M1 | Describe the non-enrolled, i.e., the participants that were selected but did not enter the study (and the reasons, if available), if applicable. |
| Drop-out | M2 | Describe the participants who dropped out from the study during the follow-up (loss to follow-up and other possible reasons: death, withdrawal, missing by design, if applicable). |
| Intermittent visit missingness | M3 | Describe the participants that have missing data for some of the measurements (intermittent, occasional omission, but do not drop out of the study). |
| Variable (item) missingness | M4 | Provide the number and proportion of missing values for each variable at each time point as appropriate for fixed or time-varying variables. Describe missingness stratifying the summaries by variables that might influence the frequency of missing values, if relevant (for example: structural variables or process variables). |
| Patterns | M5 | Describe patterns of missing values across variables at each time point and across time points. |
| Optional extensions: Missing data |  |  |
| Non-enrollment | ME1 | Compare the characteristics of the participants that entered the study with those of the non-enrolled or with the characteristics of the target population, if applicable and data are available. |
| Probability of drop-out | ME2 | Estimate the probability of drop-out after inclusion, taking appropriately into account the reasons for drop-out. |
| Dropout effect on outcome | ME3 | Visualize mean profiles of a continuous outcome by time metric stratified by time to drop-out. |
| Predictors of missingness | ME4 | Explore whether there are predictors of missingness by comparing complete vs incomplete cases or investigate predictors of time to dropout, as appropriate; this can assist in understanding of the missing data mechanism. |
| **IDA screening domain: Univariate descriptions** |  |  |
| Description of the variables at baseline | U1 | Summarize the variables used in the analysis (outcome, explanatory variables, structural variables, auxiliary variables) with numerical and graphical summaries at baseline. |
| Description of the time-varying variables at later points | U2 | Summarize the time-varying variables used in the analysis also at later time points. This might require discretization of time intervals and/or the use of different time metrics. |
| **IDA screening domain: Multivariate descriptions** |  |  |
| Association at baseline | V1 | Evaluate the association between each explanatory variable with the structural variables at baseline (with graphs and summaries). |
| Correlation at baseline | V2 | Quantify association with pairwise correlation coefficients between all explanatory variables in a matrix or heatmap at baseline. |
| Interactions at baseline, if applicable | V3 | Evaluate bivariate distributions of the variables specified in the analysis strategy with an interaction term; include appropriate graphical displays. |
| Optional extensions: Multivariate descriptions |  |  |
| Stratification | VE1 | Compute summary statistics and describe variation between strata defined based on process variables, e.g. centers, providers, locations, or by structural variables or other variables described as stratification variables in the analysis strategy (at baseline, other time points/time intervals can be also included). |
| Associations and correlations at time-points beyond baseline | VE2 | Associations and correlations between explanatory variables at time points later than baseline to explore their possible change across time; this could be useful for the identification of auxiliary variables. |
| **IDA screening domain: Longitudinal aspects** |  |  |
| Profiles | L1 | Summarize changes and variability of the outcome variable within subjects, e.g. profile plots (spaghetti-plots) for groups of individuals. |
| Trends | L2 | Describe numerically or graphically longitudinal(average) trends of the outcome variable. |
| Correlation and variability | L3 | Estimate the strength of the within-participant correlation of the outcome variable between time points and its variability across time points. |
| Trends of time-varying explanatory variables | L4 | Describe numerically or graphically the longitudinal trends of the time-varying explanatory variables. |
| Optional extensions: Longitudinal aspects |  |  |
| Cohort/Period effects | LE1 | If appropriate, summarize possible cohorts or period effects (for example, age birth cohorts or period cohorts defined by the calendar time/wave of measurement) on the outcome, and on the explanatory variables, to assess if the variation of the outcome can occur because of these effects. |

### 3.1 Participation profile

#### Aim

(1) to summarize the participation pattern of individuals in the study over time; (2) to describe the time metric(s).

Participation profile refers to temporal patterns in participation. The number of participating individuals, the number of times they were measured, and the distribution of the number of measurements per time point and per individual are described in this IDA screening domain.

Different choices of time metrics are possible depending on the research question. It can be time since inclusion in the study, time since an event, calendar time, age, or measurement occasion (defined as order of pre-planned measurement times for a participant). In some studies, it may be useful to use more than one time metric to describe the study.

Most timescales induce subject-specific times of measurements, which is naturally handled in regression analyses (for instance with mixed models that use the actual times of measurement and where using measurements at time points that are not common for all subject is not problematic), but this poses an additional challenge for summary statistics during IDA steps. When subject-specific times remain closely linked to a shared timescale, for instance planned visits or waves (nominal times), IDA can be done according to the shared timescale, with a mention of the variability the approximation in time induced. The deviations between nominal and actual times should also be explored. In other contexts, for instance when using age as the time scale in cohorts with heterogeneous ages at baseline, relevant intervals of time need to be considered for summary statistics and overall trends.

The description of the number of observations at each time point in studies with pre-planned times of observations provides information about missing values (discussed more in detail in the next domain), while it does not in study designs that foresee random times of observation [7].

### 3.2 Missing values

#### Aim

(1) to describe missing data over time and by types of missingness (non-enrollment, intermittent visit missingness, loss to follow-up, missing by design, or death); (2) to summarize the characteristics of participants with missing values over time; (3) to describe the variables with missing values (4) to find possible patterns of missing data across variables; and (5) to evaluate possible predictors of missingness and missing values.

Longitudinal data with complete information are very rare, and missing data are one of the major challenges in the design and analysis of longitudinal studies. Different analysis methods can rely on different assumptions about the missing data mechanism. An incorrect handling of missing data can lead to biased and inefficient inference [12]; therefore, a thorough investigation of the pattern of observed missingness before the beginning of the statistical analysis can have major implications for the interpretation of the results or imply possible changes in the analysis strategy.

In longitudinal studies it is important to distinguish between unit missingness (of participants) due to non-enrollment (participants that fulfill inclusion criteria that do not participate in the study), intermittent visit missingness (a missing visit) and dropout, defined as visit missingness due to attrition/loss-to-follow-up (missing values for participants that previously participated in the study); participants can also have incomplete follow-up due to death.

It is also possible that some variables (outcome and/or explanatory variables) are missing among participants for which the measurements of the other variables are available at the same visit; this type of *partial* missingness, at variable rather than participant level, is often defined as variable or item missingness. It is possible that the methods used to handle different types of missingness in the analyses differ (for example, survey weights, multiple imputation, maximum likelihood estimation), and the analysis strategy determines which aspects of missing value is important to describe.

Missing values in exploratory variables can be handled either by considering complete cases or by performing multiple imputation (MI), while the imputation of outcome in ML mixed-based models is not needed as the model intrinsically handles the missing data in the outcome. In survey studies unit non-enrollment missingness is often addressed using survey weights, which can be used to adjust the analyses for the selection of participants that makes the sample non-representative of its target population.

The number and the known characteristics of the non-responders should be described, as well as the characteristics of the participants that are lost during follow-up, the corresponding time points and reasons, if available, and the time of last observed response.

To understand how non-enrollment influences the characteristics of the available sample, some of the main characteristics of the enrolled and non-enrolled can be compared, if data are available, or the sample of enrolled can be compared to the target population. It is also useful to estimate the probability of drop-out after inclusion during study, stratifying by structural variables. The display of the mean outcome as a function of time stratified by different drop-out times can suggest a relationship between the outcome and the drop-out process [6].

For item missingness, the frequency and reasons for missing data within single explanatory variable, and the co-occurrence of missing values across different variables (for example, using visualization techniques as clustering of indicators of missing values) may be used to identify patterns of missingness. The characteristics and number of the participants for which an individual item is missing can also be described separately.

Predictors of missing values can be identified by comparing the characteristics of subjects with complete and incomplete data at each measurement occasion; it is common to compare the baseline characteristics, where the extent of missing values is usually smaller compared to longitudinal measurements.

Another aim within this domain can be to identify potential auxiliary variables, i.e., variables not required for the analysis but that can be used to recover some missing information through their correlation with the incomplete variables, for example via inclusion in an imputation model (if envisioned in the analysis strategy) or for the construction of survey weights. As this often requires looking at the correlation between variables, this can be assessed via the multivariate descriptions.

### 3.3 Univariate descriptions

#### Aim

(1) to describe all variables that are used in the analysis (outcomes, explanatory variables, structural variables, auxiliary variables) with numerical and graphical summaries at baseline; (2) to describe the time-varying variables at all time points.

The univariate descriptions explore the characteristics of the variables, one at a time. The results can be used to evaluate if the observed distributions are as expected, or to identify problematic aspects (unexpected values, sparse categories, etc). Descriptive statistics can be used to summarize the variables, as described in [4].

The time-varying variables should be summarized also at time points after baseline. As evoked earlier, discretization into intervals may be indicated if the time metric is on a continuous rather than on a categorical scale and the number of different observed times is large. Different time metrics can be used to summarize the variables. Using the time metric of the data collection process can be useful for the identification of data collection problems (e.g., specific characteristics or problems in some waves). In contrast, the time metric linked to the analysis strategy can provide more useful information about the distributions of the variables to be modelled.

### 3.4 Multivariate descriptions

#### Aim

(1) to provide summaries of the explanatory variables stratified by structural variables or by process variables (e.g., variables that describe the process under which data was collected, might be centers, providers, locations); (2) to describe associations and correlations between explanatory variables (focusing mostly on baseline values); (3) to provide stratified summaries of the data.

The explorations proposed in the multivariate domain are very similar to those proposed in the context of IDA for regression modeling [4], and include the exploration of associations between exploratory variables with structural variables, and the evaluations of associations and correlations among exploratory variables. If interactions between explanatory variables are considered, the exploration of the association between these variables should be carefully addressed in IDA [4].

We suggest to focus primarily on associations between variables at baseline (where usually the missing values are less common). Follow-up times can be considered if the aim is to evaluate if/how the associations and correlations change during follow-up; however, the interpretations should be cautious, as the results are based only on observed data and the missing data mechanism that occurs during follow-up can alter the associations.

The distributions of explanatory variables stratified by the values of the structural variables are also described in the multivariate descriptions; the considerations about the influence of missing values on the results apply also for these descriptions; numerical structural variables might require some type of discretization.

### 3.5 Longitudinal aspects

#### Aim

(1) to describe longitudinal trends of the time-varying variables including changes and variability within and between subjects; (2) to evaluate the strength of correlation of the repeated measurements across time points.

The exploration of the characteristics of the participants through time is of upmost importance and should be described using the time metric chosen in the analysis strategy. The repeated measurements from the same subject in longitudinal studies are usually correlated, thus IDA should explore the trend of the repeated variables but also the degree of dependence within subjects by evaluating the variance, covariance and correlation on repeated measurements of the outcome variable.

The time-varying explanatory variables can be explored; these explorations are useful for providing domain experts a description of some of the characteristics of the sample that can be compared to the expected. As discussed earlier, descriptive summaries based on the observed longitudinal data might be biased, and should therefore be interpreted carefully.

In many applications it is important to summarize the cohort (individuals who experience the same event in the same time) or period (time when the participants are measured) effect on the outcome and on the exploratory variables. The design of the longitudinal study might make the effect of age, cohort and period difficult to separate and subject to confounding. The results from IDA explorations might indicate the need to take cohort or period effects into account in the modelling.

## 4 Case study: Age-associated decline in grip strength in the Danish data from the SHARE study

To illustrate the use of the data screening checklist for longitudinal data we conducted the IDA screening step for a case study, where the research aim was to evaluate the age-associated decline in grip strength. An IDA plan was developed (Supplementary file 1) and a reproducible and structured IDA report for the analysis was implemented using R language [13] (version 4.0.2) and made available at https://stratosida.github.io/longitudinal/; the report presents the full IDA data screening results and provides the R code for reproducibility.

Firstly, we briefly illustrate the data and the analysis strategy and present the IDA plan; a selected set of IDA explorations are presented in the results, and the possible consequences of the IDA findings are reported and discussed section 4.5.

### 4.1 SHARE data

We used the data from the Survey of Health, Ageing and Retirement in Europe (SHARE). SHARE is a multinational panel data survey, collecting data on medical, economic and social characteristics of about 140,000 unique participants after age 50 years, from 28 European countries and Israel [11]. The SHARE study contains health, lifestyle, and socioeconomic data at individual and household level. These data have been collected over several waves since 2004, using questionnaires and performing a limited number of performance measurements. The baseline and the longitudinal questionnaires differ in some aspects, and some questions were modified during the course of the study; in wave 3 and partly in wave 7 a different questionnaire (retrospective SHARELIFE) was used to collect retrospective information about participants. Leveraging these data for research purposes can be daunting due to the complex structure of the longitudinal design with refresher samples organized in 25 modules with about 1000 questions. Functions written in the R language [13] are available that facilitate data extraction and data preparation of SHARE data [8].

We provide an explanation and elaboration of an IDA checklist for data screening using SHARE data collected during the first seven waves 2004 to 2017 in Denmark, which based the selection of participants on simple random sampling.

### 4.2 Study aims and corresponding analysis strategy

The research question aims at assessing the age-associated decline of hand grip strength by sex, after adjusting for a set of explanatory variables that are known to be associated with the outcome (weight, height, education level, physical activity and smoking). Here we give a basic overview of the corresponding statistical analysis strategy.

The study population are individuals from Denmark aged 50 or older at first interview. The outcome is maximum grip strength measured at different interviews (recorded with a hand-held dynamometer, assessed as the maximum score out of two measurements per hand). The time metric is the age at interview. The time-fixed variables evaluated at first interview are sex, height and education (categorized in three levels); the time-varying variables are weight, physical activity (vigorous or low intensity, both dichotomized) and smoking status. Interaction terms between age and all the time-fixed variables (sex, education, height) will be included in the prespecified statistical analysis models to evaluate the association between these time-fixed variables with the trajectory of the outcome; the main interest is in the interpretation of the interaction terms between sex and functions of age on the grip strength. Nonlinear functional forms for continuous variables will be assessed using linear, quadratic, and cubic polynomials.

A linear mixed model [14] is planned to be used to address the research question. The trajectory over time of the outcome is explained at the population level using fixed effects and individual-specific deviations from the population trajectory are captured using random effects to account for the intra-individual serial correlation. The model accommodates individual-specific times of outcome measurements.

The linear mixed model, estimated by maximum likelihood, is robust to missing at random outcome data, that is when the missingness can be predicted by the observations (outcome and explanatory). Missing data at variable/item level (for the time-fixed explanatory variables) can be handled either by considering complete cases or by performing multiple imputation. We will use data from the SHARE study that are publicly available upon registration for use for research purposes (http://www.share-project.org/data-access.html). All analyses will be carried out using R statistical language [13].

### 4.3 IDA plan

The detailed IDA data screening plan for this study is described in the Supplementary file 1; it includes most of the points included in our checklist, describing the specific explorations that should be addressed and their aim.

Structural variables in the context of IDA are: sex and grouped age (because of their known association with the outcome), wave and type of interview (baseline vs. longitudinal) (because of differences in data collection process).

### 4.4 Results of IDA

Here we present the main IDA findings for each domain; the consequences are discussed in the next section.

#### 4.4.1 Participation profile

The interviews were carried out between April 2004 and October 2017, in seven Waves (Fig. 1). The median time between interviews in successive waves was about 2 years, the longest times passed between Wave 1 and 2 (median: 2.5 years, see the accompanying web site for more details).

**Fig 1.**
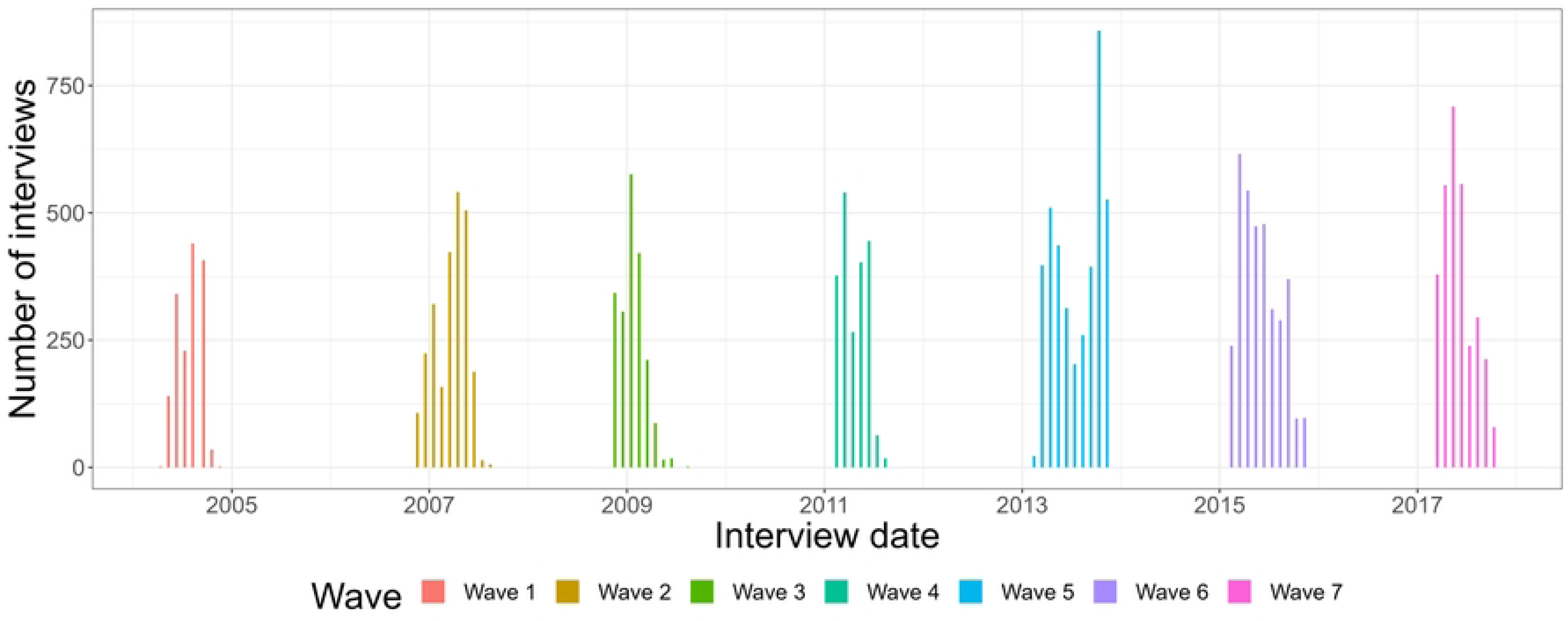
Distribution of the number of interviews carried out in Denmark in the SHARE study in time.

Overall, 5452 unique participants were interviewed 18632 times during the study. The number of participants who attended the interview in each wave, stratified by baseline wave are shown in Fig. 2, which highlights that new participants (refreshment samples) were included during the study and that Wave 5 had the most interviews. The exploration of the age at inclusion shows that full range refreshment samples were used in Wave 2 and 5, and refreshment samples only of the younger people in Wave 4 and 6 (Fig. 3), as described in the study protocol.

**Fig 2.**
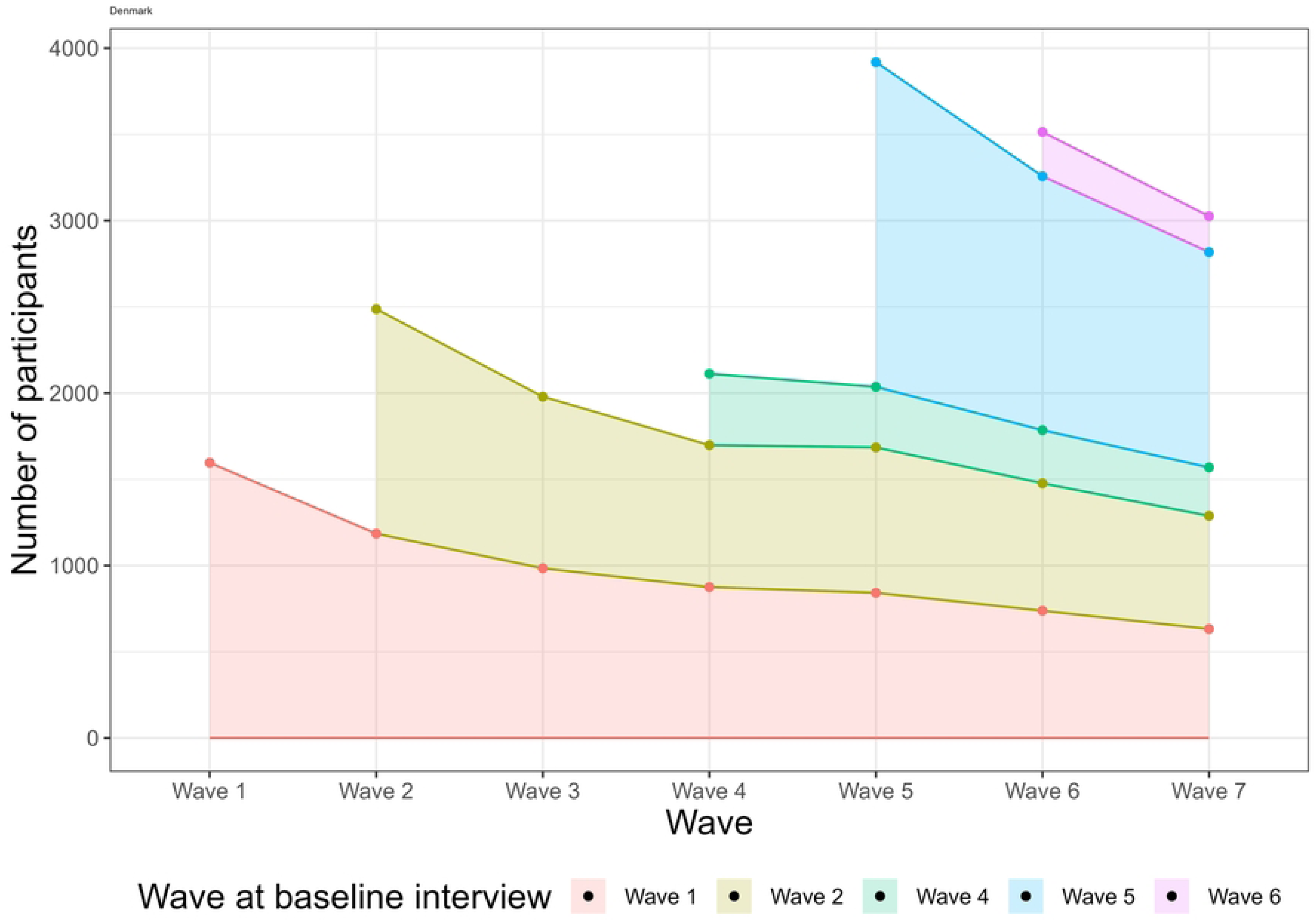
Number of participants in each wave, stratified by baseline wave.

**Fig 3.**
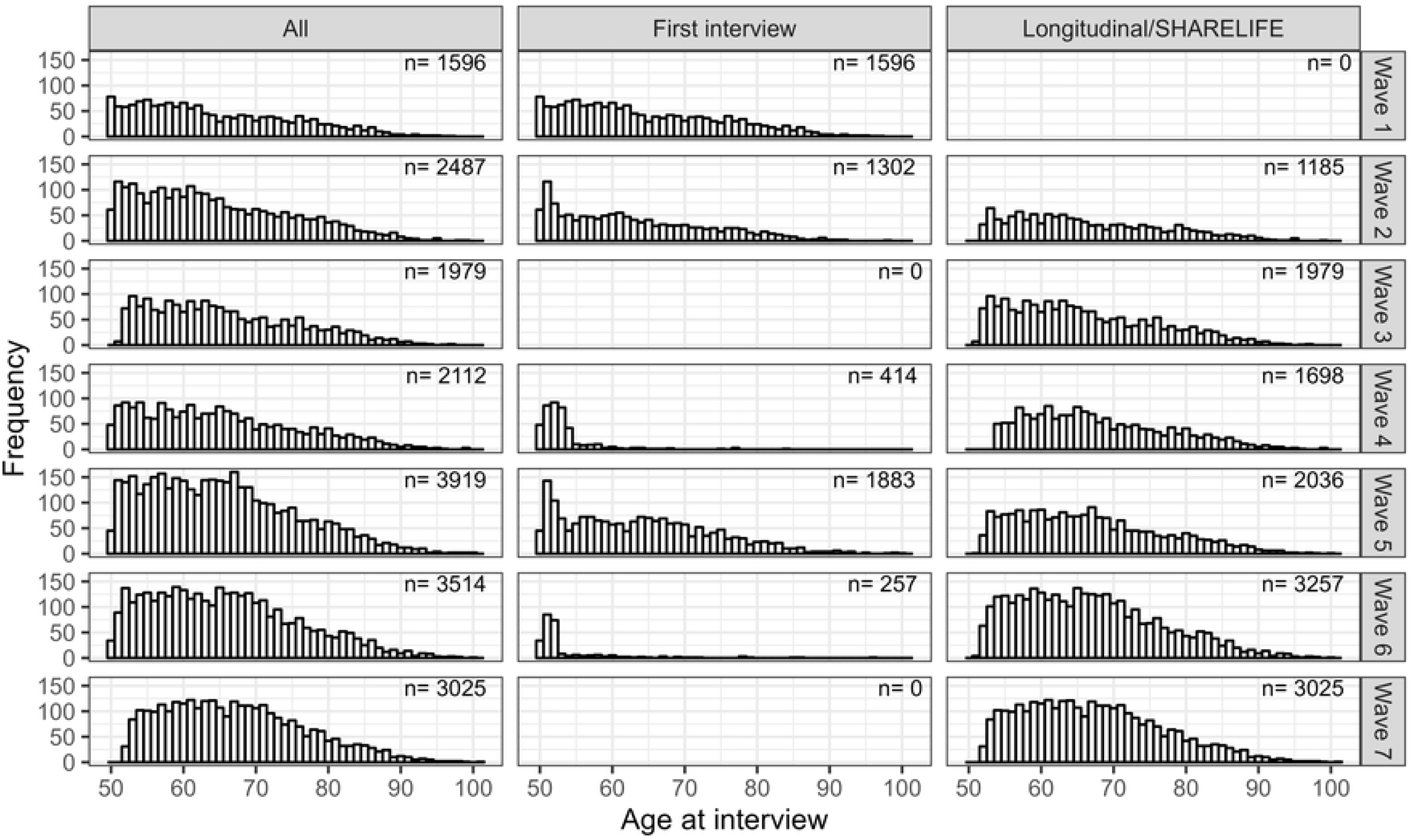
Distribution of age across waves and by baseline or longitudinal/SHARELIFE interview. Note that SHARELIFE interviews were conducted in Waves 3 (all participants) and 7 (60% of the participants).

The median and modal number of interviews per participant was 3, 18% were interviewed only once, only 22% were interviewed 6 or 7 times (Table 2); further aspects about drop-out are discussed in the missing value domain.

**Table 2.** Number of interviews per participant.

| Interviews per participant | 1 | 2 | 3 | 4 | 5 | 6 | 7 |
| --- | --- | --- | --- | --- | --- | --- | --- |
| Frequency | 965 | 966 | 1508 | 527 | 307 | 685 | 494 |
| Proportion | 0.18 | 0.18 | 0.28 | 0.10 | 0.06 | 0.13 | 0.09 |

Age is the time metric of interest in the analysis described in the analysis strategy, therefore its distribution is described in the participation profile. In later waves the participants were on average older (for example, the median age increased from 62 to 66 from Wave 1 to Wave 7), but the age distribution in the sample and in the target population was similar. Fig. 3 shows the distribution of age over waves, overall and by type of interview.

**Fig 4.**
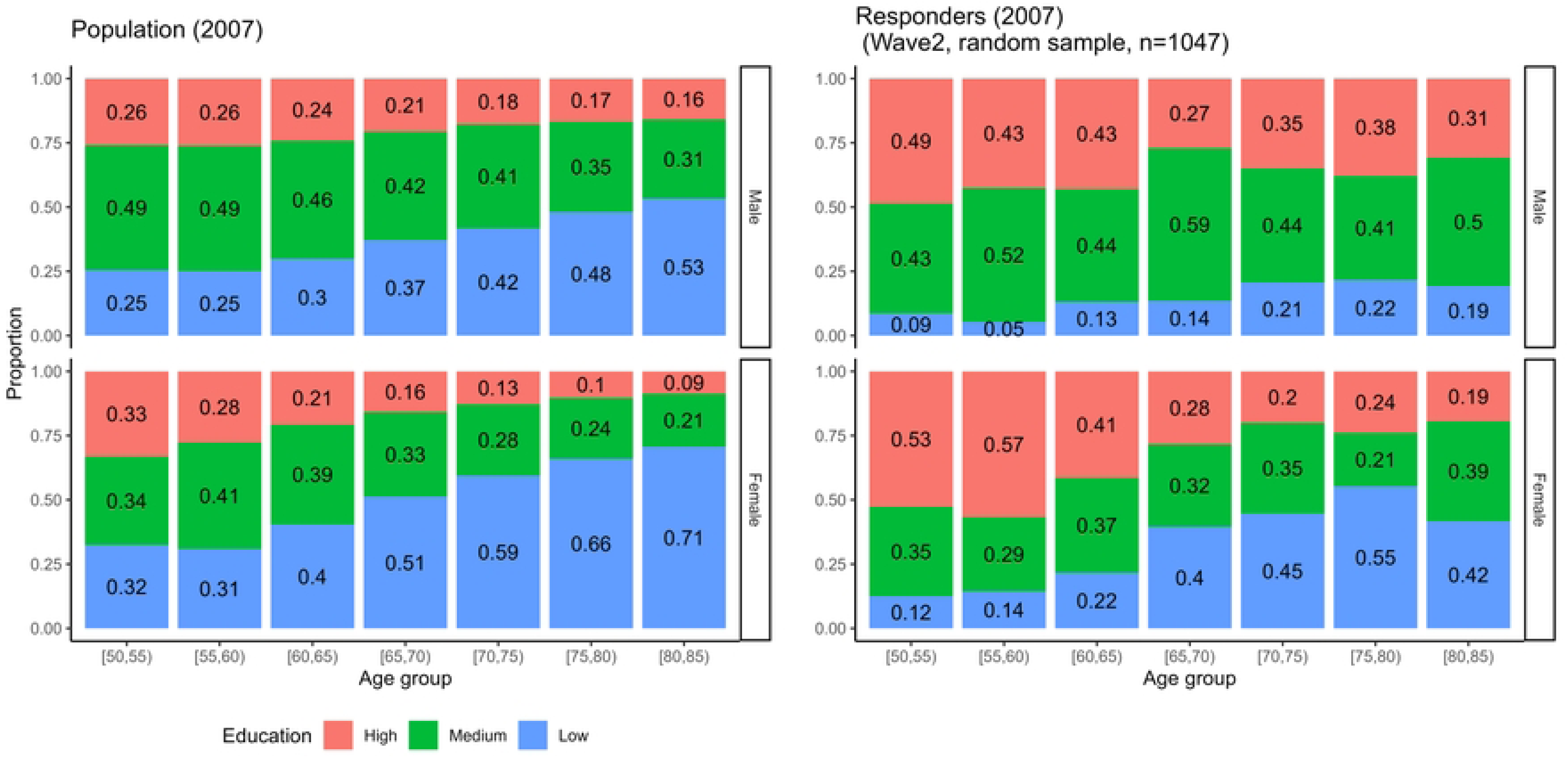
Distribution of education in the population in year 2007 in Denmark and the refreshment sample of Wave 2, by sex and age group. The analyses were limited to the ages between 50 and 85, as population data on education were unavailable for older inhabitants; the sample displayed from Wave 2 is a random sample used in this wave as refreshment sample; details are given in the online IDA report.

**Fig 5.**
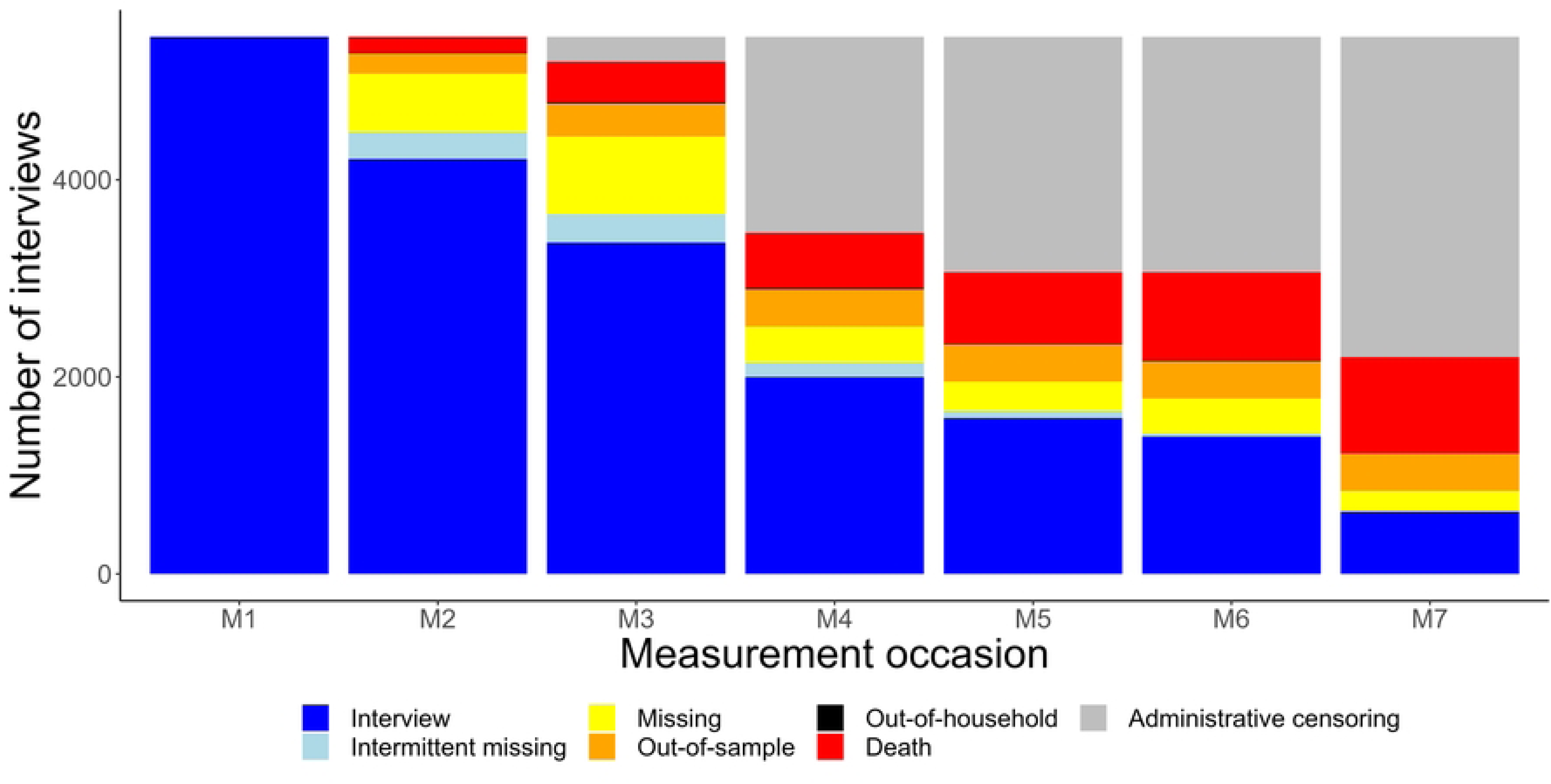
Number of participants with observed and missing data by measurement occasion and by type of missingness. Interview: participant participated with a valid interview; intermittent missingness: missing at measurement occasion but with valid interview later; missing: missing at measurement occasion and no interview later; out-of-sample: was removed from the sample because lost to follow-up (by study definition after at least three missing interviews, here the definition was applied retrospectively); out-of-household: not interviewed because not member of the household; death: died at measurement occasion or earlier; administrative censoring: did not have interview because the study ended.

The participation profile highlighted the complexity of the study design and the fact that most participants were measured few times; it also provided information about the distribution of age, which is the continuous time of interest and for which we did not identify any specific problems.

#### 4.4.2 Missing data

The characteristics of non-enrolled could be studied only through the comparison of the observed samples with some known characteristics of the target population (sex, age and education composition, data were available from the statistical office of the European Union - Eurostat https://commission.europa.eu/index_en, from year 2007, Wave 2 of the study). The aim of this comparison is to evaluate if the sample differs from the target population.

The responders that participated in the survey at least once had substantially higher education compared to the population in the same age and sex groups, males in the younger age groups were slightly underrepresented, as were older females (Fig. **??** for the distribution of education for data from Wave 2, the complete results are similar for the other waves and presented in the online IDA report).

Many participants that entered the study had missing data during the longitudinal follow-up. The deaths were reported with high quality and timely, as only 1% of the participants had unknown vital status at the end of the study and overall, 978 deaths were reported by Wave 7.

In Fig. 6 participants were classified in seven categories based on their participation at each measurement occasion (defined as number of waves since first measurement). The figure highlights that some participants had intermittent missingness, missingness by design because participants were not eligible (out-of-household) was very rare, while administrative censoring was common due to the study design (for example, many new participants were included in Wave 5 and the follow-up ended in Wave 7), and so were deaths and losses to follow-up (missing and out-of-sample).

**Fig 6.**
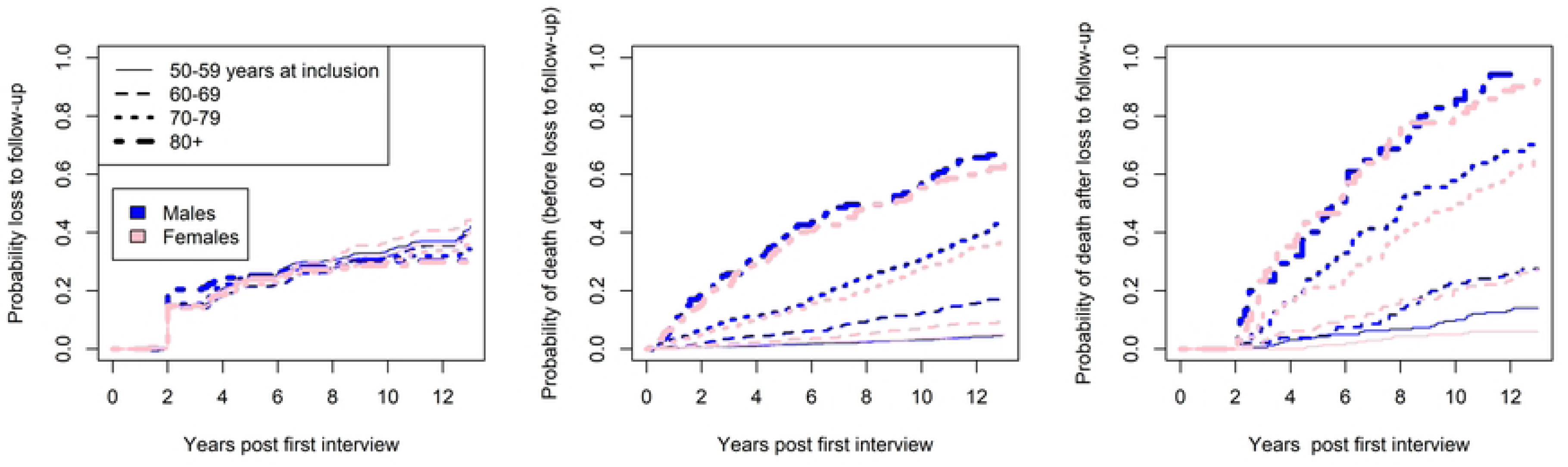
Cumulative incidence estimates of loss to follow-up, death without loss to follow-up and death after loss to follow-up, stratified by sex and age category. The first two incidence functions are obtained using the Aalen-Johansen estimator, the third is based on the Kaplan-Maier estimator. Aalen-Johansen estimators, stratified by sex and age group at first interview, obtained using the survival R package.

For the analysis purposes, the participants of some of the groups described in Fig. 6 would be classified as lost to follow-up (out-of-sample, missingness, out-of-household if not re-included in the sample later); using this definition we estimated the probability of loss to follow-up, death and death after follow-up. Estimate of cumulative incidence functions (using Aalen-Johansen estimators for loss to follow-up and deaths) indicated that the probability of loss to follow-up was virtually the same across age groups and sex. In contrast, the probability of death prior and post loss to follow-up substantially increased with age as expected, and tended to be higher for males at younger ages (Fig. 6). Additional analyses showed that participants that died differed from the others also because they were more frequently smokers, had lower education and engaged in less physical activity, and had considerably lower levels of grip strength at baseline measurement (online IDA report); compared to complete responders, those that dropped out of the study for reasons different than death, had lower education, less physically activity and smoked more frequently (online IDA report).

The mean outcome profiles of participants that died during follow-up were lower compared to those that survived, especially among older males (Fig. 7, left panel), while the difference in outcome between complete and incomplete cases due to loss to follow-up was smaller (Fig. 7, right panel).

**Fig 7.**
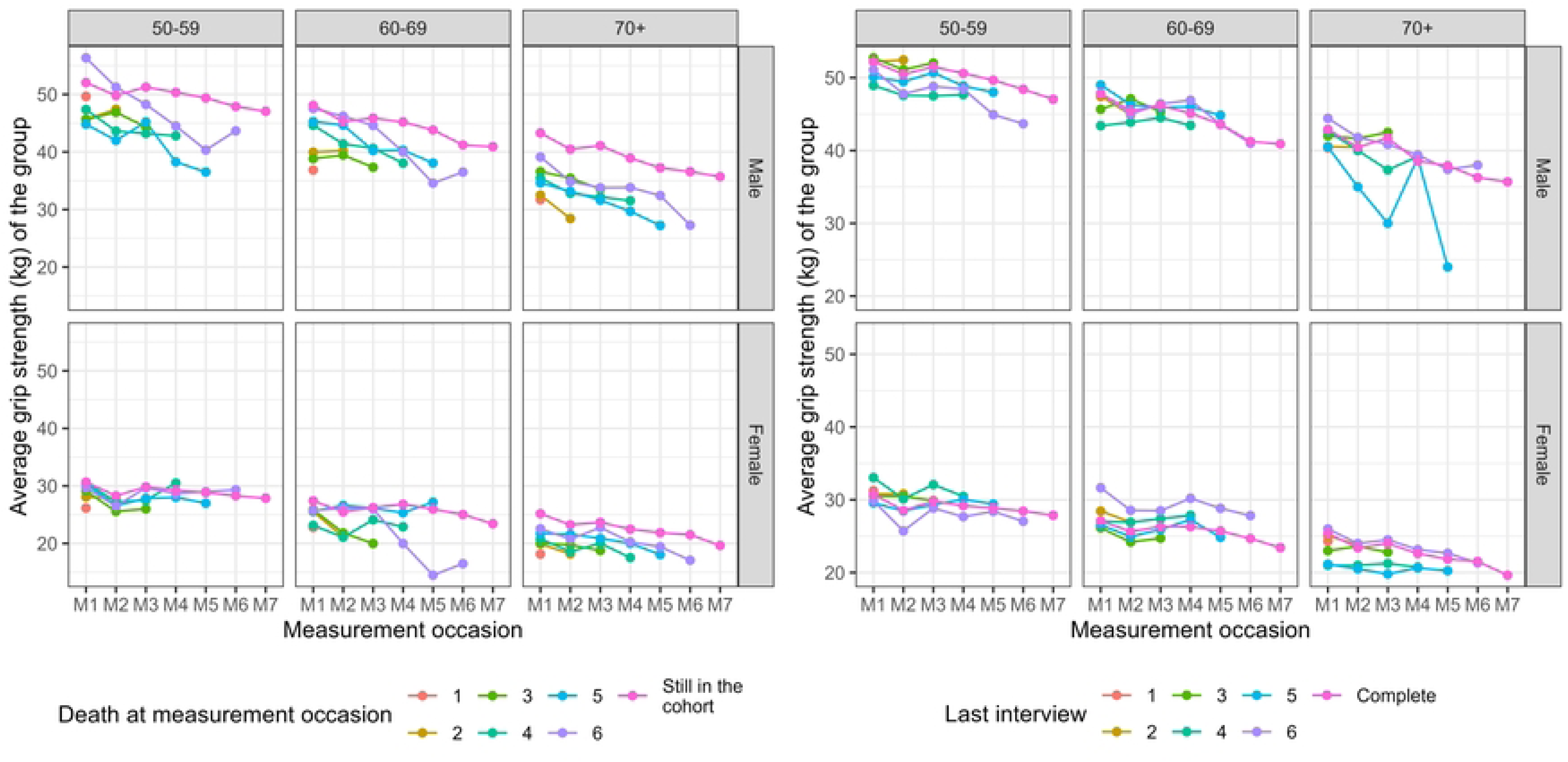
Mean maximum grip strength for groups with reported death (left panel) or with loss to follow-up (right panel) at different measurement occasions, stratified by age group and sex. Participants classified in the groups still in the cohort had complete measurements for 7 waves.

We explored the amount of missing outcomes among the interviews that were conducted (item missingness in the outcome) to evaluate the frequency of outcome missingness with valid interview, and its association with the characteristics of the participants. The amount of this type of outcome missingness varied from 2.2 to 6.5% across measurement occasions, females had more missing values than males and the proportion of missingness increased with longer follow-up (Table 4) and with age (Table 3). Participants with missing outcome were unable to take the measurement in 36% of cases, indicating that missing values might be related to bad physical conditions; 21% refused to take the measurement, 2% had a proxy interview, while the reason for missingness was unknown for the others.

**Table 3.** **Percentage (%) and number (n) of missing values in the outcome (maximum grip strength) among participants that were interviewed,** by age group and sex using all available data.

|  | 50-59 | 60-69 | 70-79 | 80+ |
| --- | --- | --- | --- | --- |
| **Males** |  |  |  |  |
| % missing | 1.5 | 1.9 | 3.1 | 11.4 |
| n/Total | 45/2890 | 57/2989 | 63/1994 | 79/611 |
| **Females** |  |  |  |  |
| % missing | 2.4 | 2.7 | 6.2 | 13.8 |
| n/Total | 77/3159 | 89/3226 | 140/2104 | 153/956 |

**Table 4.** Percentage (%) and number (n) of missing values in the explanatory variables and outcome by measurement occasion and sex. PA: physical activity. Here we show only the first interview data for variables used as time-fixed in the model (height, education and smoking - following the change suggested by IDA) and remove the observations missing by design.

|  | M1 | M2 | M3 | M4 | M5 | M6 | M7 |
| --- | --- | --- | --- | --- | --- | --- | --- |
| **Weight** |  |  |  |  |  |  |  |
| Males, % | 0.70 | 0.65 | 0.63 | 0.96 | 0.83 | 1.24 | 1.02 |
| n | 18/2583 | 10/1528 | 7/1107 | 9/940 | 6/720 | 8/646 | 3/294 |
| Females, % | 2.54 | 2.25 | 3.54 | 3.21 | 2.56 | 2.27 | 2.07 |
| n | 73/2869 | 38/1688 | 45/1272 | 34/1059 | 22/861 | 17/748 | 7/338 |
| **PA vigorous** |  |  |  |  |  |  |  |
| Males, % | 0.66 | 1.20 | 0.57 | 1.01 | 0.28 | 0.48 | 0.00 |
| n | 17/2583 | 17/1419 | 3/526 | 8/793 | 2/720 | 3/630 | 0/257 |
| Females, % | 0.42 | 1.57 | 1.98 | 1.30 | 0.00 | 0.00 | 0.00 |
| n | 12/2869 | 25/1589 | 12/605 | 12/925 | 0/861 | 0/732 | 0/309 |
| **PA moderate** |  |  |  |  |  |  |  |
| Males, % | 0.66 | 1.20 | 0.57 | 1.01 | 0.28 | 0.32 | 0.00 |
| n | 17/2583 | 17/1419 | 3/526 | 8/793 | 2/720 | 2/630 | 0/257 |
| Females, % | 0.45 | 1.57 | 1.98 | 1.30 | 0.00 | 0.00 | 0.00 |
| n | 13/2869 | 25/1589 | 12/605 | 12/925 | 0/861 | 0/732 | 0/309 |
| **Grip strength** |  |  |  |  |  |  |  |
| Males, % | 2.75 | 2.02 | 2.50 | 2.87 | 3.19 | 4.49 | 5.10 |
| n | 71/2583 | 40/1983 | 39/1562 | 27/940 | 23/720 | 29/646 | 15/294 |
| Females, % | 3.80 | 3.68 | 5.16 | 5.38 | 5.11 | 6.02 | 8.58 |
| n | 109/2869 | 82/2228 | 93/1801 | 57/1059 | 44/861 | 45/748 | 29/338 |
| **Time-fixed variables at baseline** |  |  |  |  |  |  |  |
|  | **Height** | **Education** | **Smoking** |  |  |  |  |
| Males, % | 0.55 | 0.54 | 0.62 |  |  |  |  |
| n | 13/2583 | 14/2583 | 16/2583 |  |  |  |  |
| Females, % | 0.73 | 0.35 | 0.45 |  |  |  |  |
| n | 21/2869 | 10/2869 | 10/2869 |  |  |  |  |

There was no clear association between missingness in different measurement occasions in the outcome, and a relatively small proportion of subjects had outcome missingness in more than one occasion, when the interview was performed (Fig. 8).

**Fig 8.**
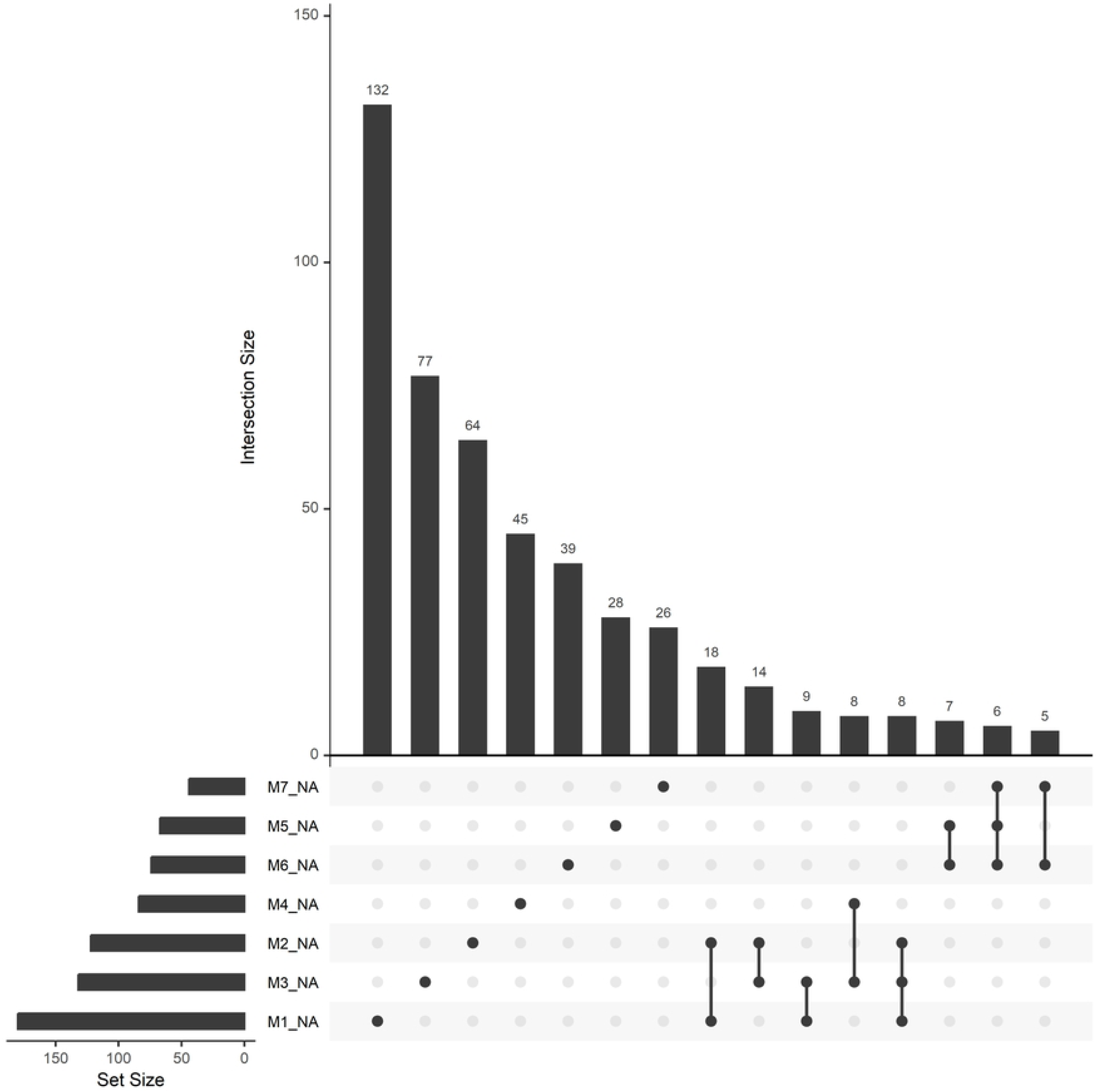
Co-occurrence of outcome missingness across measurement occasions. The number on the bars indicate the number of participants that have certain variables missing together (the missing variables are indicated using dots on the horizontal axis, M1 NA indicates that the variable is missing at first measurement occasion, etc.).

In this case study, item missingness of the explanatory variables is considered separately from unit missingness, as the analysis strategy considers using multiple imputations to handle item missingness of the explanatory variables, or complete case analysis if the amount of missing values is relatively small.

Some of the time-varying explanatory variables were missing by design (weight in Wave 3 and physical activity variables in SHARELIFE interviews, current smoking in longitudinal interviews in Waves 6 and 7), as highlighted by Fig. 9. The analyst might thus decide to consider smoking status at baseline rather than current smoking in the statistical analysis. Item missingness was very low for all variables when missing by design missingness was not considered (Table 4).

**Fig 9.**
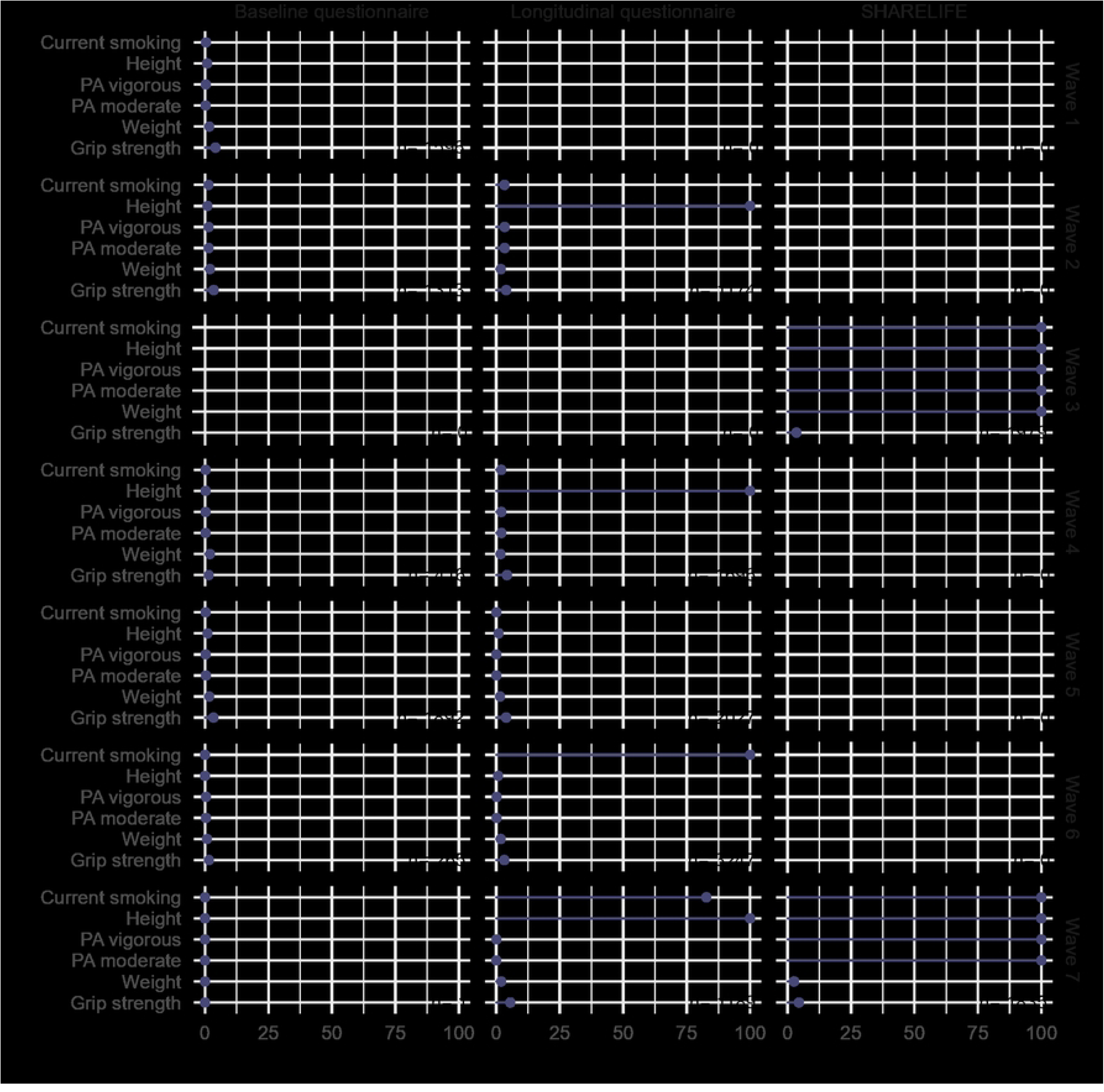
Graphical representation of the percentage of missing values (item missingness) for time varying variables, stratified by wave and type of interview and for the outcome. By design new participants were not included in Wave 3 or 7, SHARELIFE interviews were conducted in Wave 3 (all participants) and in partly in Wave 7 (only for participants that did not have a SHARELIFE interview in Wave 3, about 60%). n is the sample size.

#### 4.4.3 Univariate descriptions

The characteristics of the participants at baseline interview are summarized in Table 5 (overall and by sex, discussed in the multivariate descriptions). The summary statistics did not indicate specific problems (unexpected location or variability values for numerical variables, sparse categories for categorical variables).

**Table 5.** Descriptive statistics of the baseline characteristics of *n* = 5452 participants, overall and stratified by sex. Md (Q1, Q3) represent the median, lower quartile and the upper quartile for continuous variables. Numbers after percentages are frequencies.

|  | Non-missing | Overall<br>(n=5452) | Male<br>(n=2583) | Female<br>(n=2869) |
| --- | --- | --- | --- | --- |
| Sex : Female | 5452 | 53% (2869) |  |  |
| Age Md (Q1, Q3) | 5452 | 60 (53, 70) | 60 (53, 69) | 60 (53, 70) |
| Age groups : 50-59 | 5452 | 47% (2576) | 48% (1230) | 47% (1346) |
| 60-69 |  | 28% (1502) | 28% (734) | 27% (768) |
| 70-80 |  | 19% (1012) | 18% (472) | 19% (540) |
| 80+ |  | 7% (362) | 6% (147) | 7% (215) |
| Education : Low | 5428 | 22% (1191) | 15% (397) | 28% (794) |
| Medium |  | 39% (2130) | 47% (1204) | 32% (926) |
| High |  | 39% (2107) | 38% (968) | 40% (1139) |
| Weight (kg) | 5361 | 75 (65, 85) | 82 (75, 92) | 68 (60, 77) |
| Height (cm) | 5418 | 171 (165, 178) | 178 (173, 183) | 165 (161, 170) |
| Vigorous PA | 5423 | 60% (3274) | 64% (1632) | 57% (1642) |
| Low intensity PA | 5422 | 90% (4886) | 91% (2331) | 89% (2555) |
| Current smoking : Yes | 5423 | 26% (1395) | 27% (696) | 24% (699) |

The variables weight, height and grip strength were reported with terminal digit preference (values ending with 0 and 5 were more frequent than expected). Fig. 10 shows the distribution of grip strength and indicates that digit preferences did occur with examiners choosing more likely numbers ending with 0 or 5. This likely increases measurement error, and the IDA suggests that the impact on regression analyses would be worth exploring. The bimodality of the distribution is due to the inclusion of males and females, as shown in multivariate descriptions.

**Fig 10.**
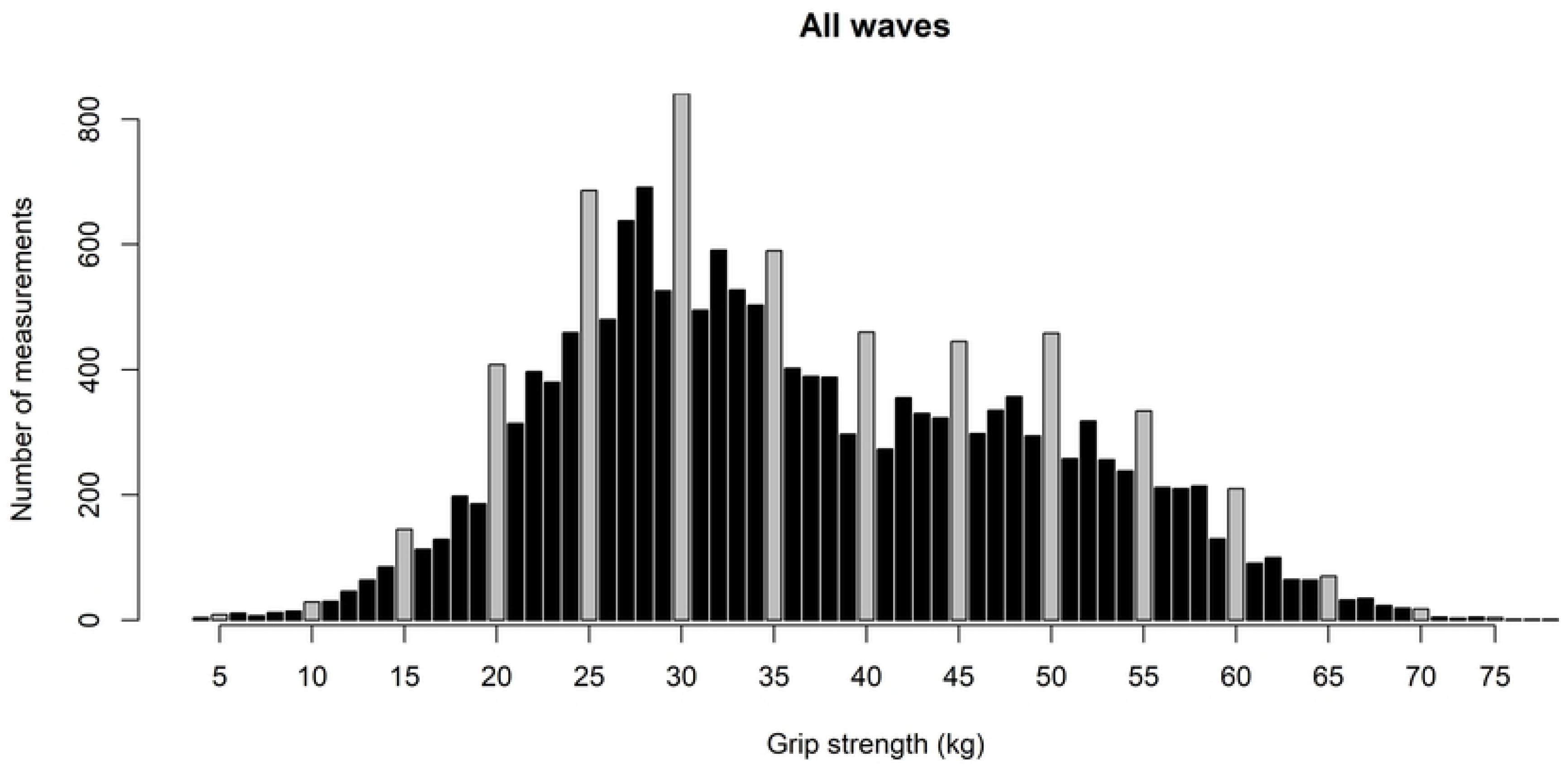
Distribution of maximum grip strength across all participants. (gray bars indicate numbers ending with figure 0 or 5).

#### 4.4.4 Multivariate descriptions

Sex is a structural variable in our case study, therefore the distributions of all the explanatory variables, stratified by sex, are explored in the multivariate descriptions. Females and males differed substantially in the distribution of height, weight, vigorous (but not low-intensity) physical activity, and education, while the distribution of age and the proportion of current smokers was similar (Supplementary file 2 and accompanying web site).

The bimodal distribution of grip strength was explained by the large average differences between males and females and the histogram of age indicated that a Gaussian distribution assumption at each wave is appropriate when separated by sex (Fig. 11).

**Fig 11.**
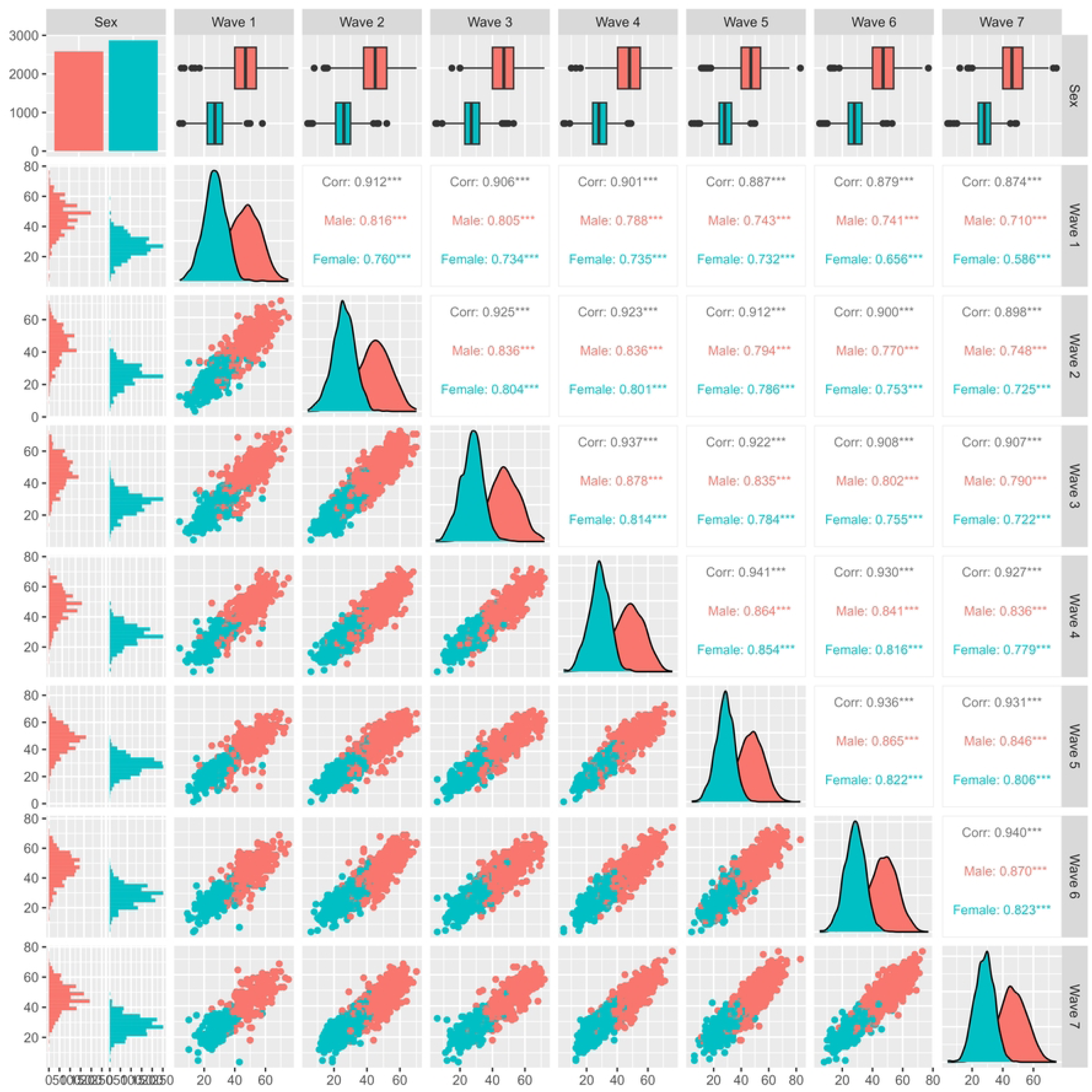
Generalized pairs plot for grip strength, across waves and by sex.

As expected, at baseline the couples of variables with highest positive correlation were weight and height, and the two variables measuring physical activity (Table 6 for overall correlations and Fig. 12 stratifying by sex).

**Fig 12.**
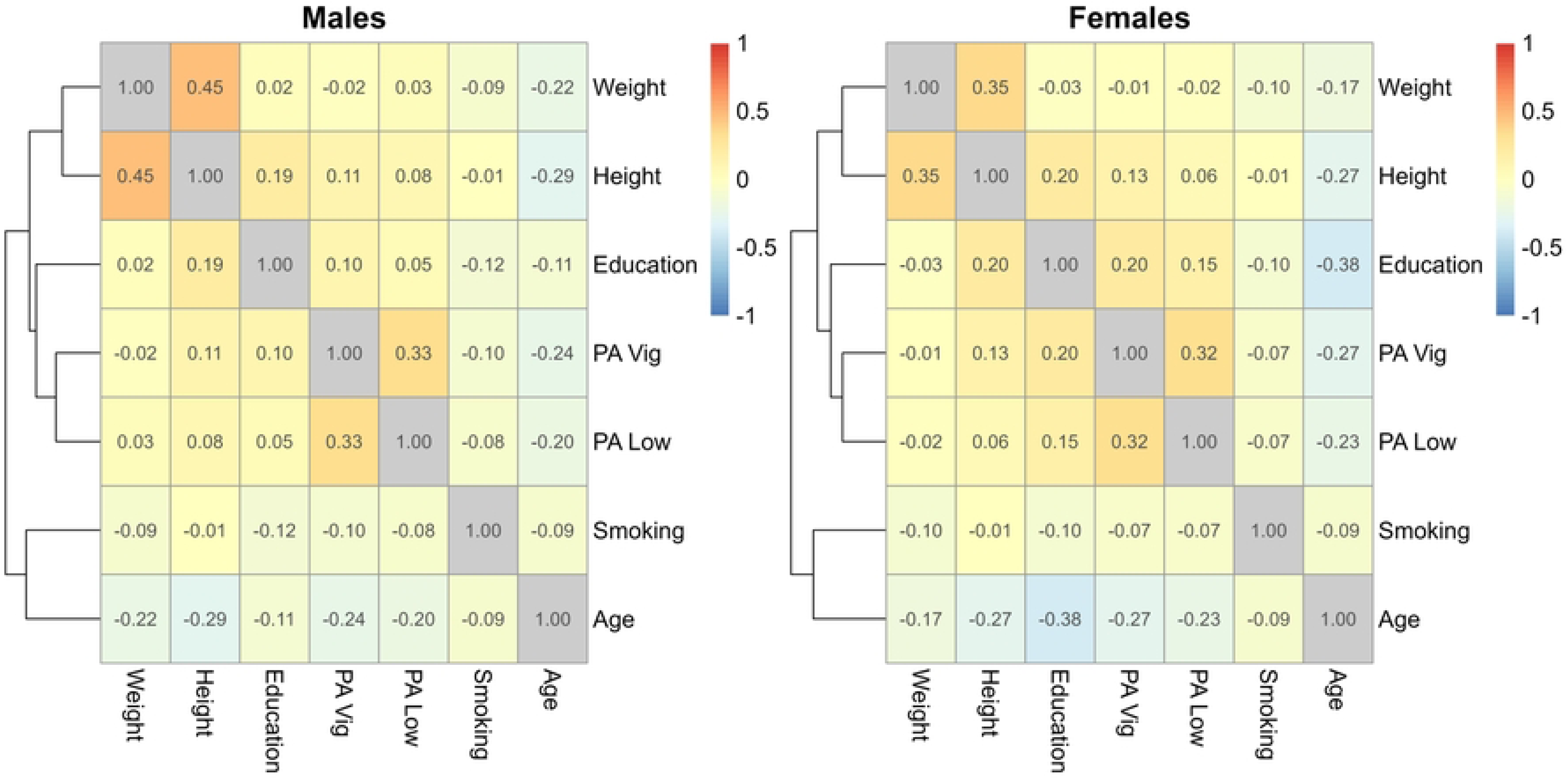
Correlation between explanatory variables at baseline, stratified by sex; the education levels were used as numbers.

**Table 6.**
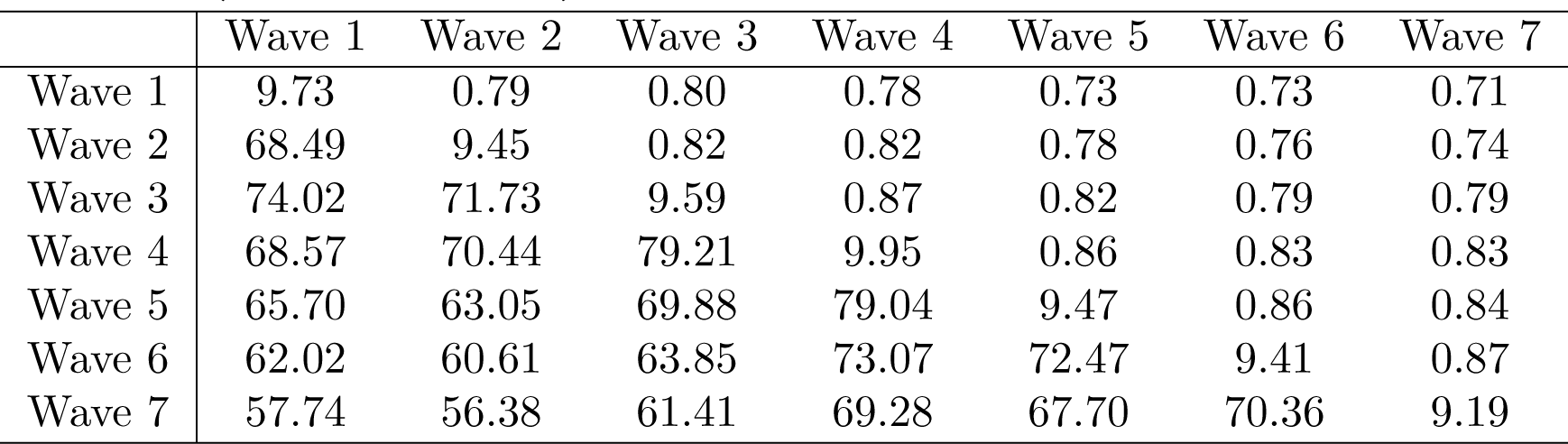
Correlations (above diagonal), standard deviations (diagonal) and covariances (below diagonal) of grip strength across waves for males.

|  | Wave 1 | Wave 2 | Wave 3 | Wave 4 | Wave 5 | Wave 6 | Wave 7 |
| --- | --- | --- | --- | --- | --- | --- | --- |
| Wave 1 | 9.73 | 0.79 | 0.80 | 0.78 | 0.73 | 0.73 | 0.71 |
| Wave 2 | 68.49 | 9.45 | 0.82 | 0.82 | 0.78 | 0.76 | 0.74 |
| Wave 3 | 74.02 | 71.73 | 9.59 | 0.87 | 0.82 | 0.79 | 0.79 |
| Wave 4 | 68.57 | 70.44 | 79.21 | 9.95 | 0.86 | 0.83 | 0.83 |
| Wave 5 | 65.70 | 63.05 | 69.88 | 79.04 | 9.47 | 0.86 | 0.84 |
| Wave 6 | 62.02 | 60.61 | 63.85 | 73.07 | 72.47 | 9.41 | 0.87 |
| Wave 7 | 57.74 | 56.38 | 61.41 | 69.28 | 67.70 | 70.36 | 9.19 |

Age was negatively correlated with all the explanatory variables, the negative association between age and education among females was the highest, which can be explained by the study design, where there is a strong association between age and birth year. The SHARE study encompasses multiple age cohorts, followed in different calendar periods, as summarized in Supplementary file 2 and as expected due to the design of the cohort.

#### 4.4.5 Longitudinal aspects

To visualize individuals’ grip strength trajectories we used profile plots; interactive plots are also available (online IDA report). The profiles based on subgroups of participants facilitate the visualizations of individual trajectories (Fig. 13 and Supplementary file 2 show 100 random participants per group of initial grip strength quantile), which are not visible using complete data when the number of participants is large (Fig. 14 and Supplementary file 2 show the profiles for all data). Even though age was included as a continuous time metric in the analysis strategy, a summary stratified by ten-year groups can serve as a quick overview of the longitudinal trends by age. The graphs that use age as a time metric give an idea of the shape of trajectory for model specification (which has to be determined a priori), those based on measurement occasion give a clearer overview of the individual trajectories, as participants enter the study at different ages.

**Fig 13.**
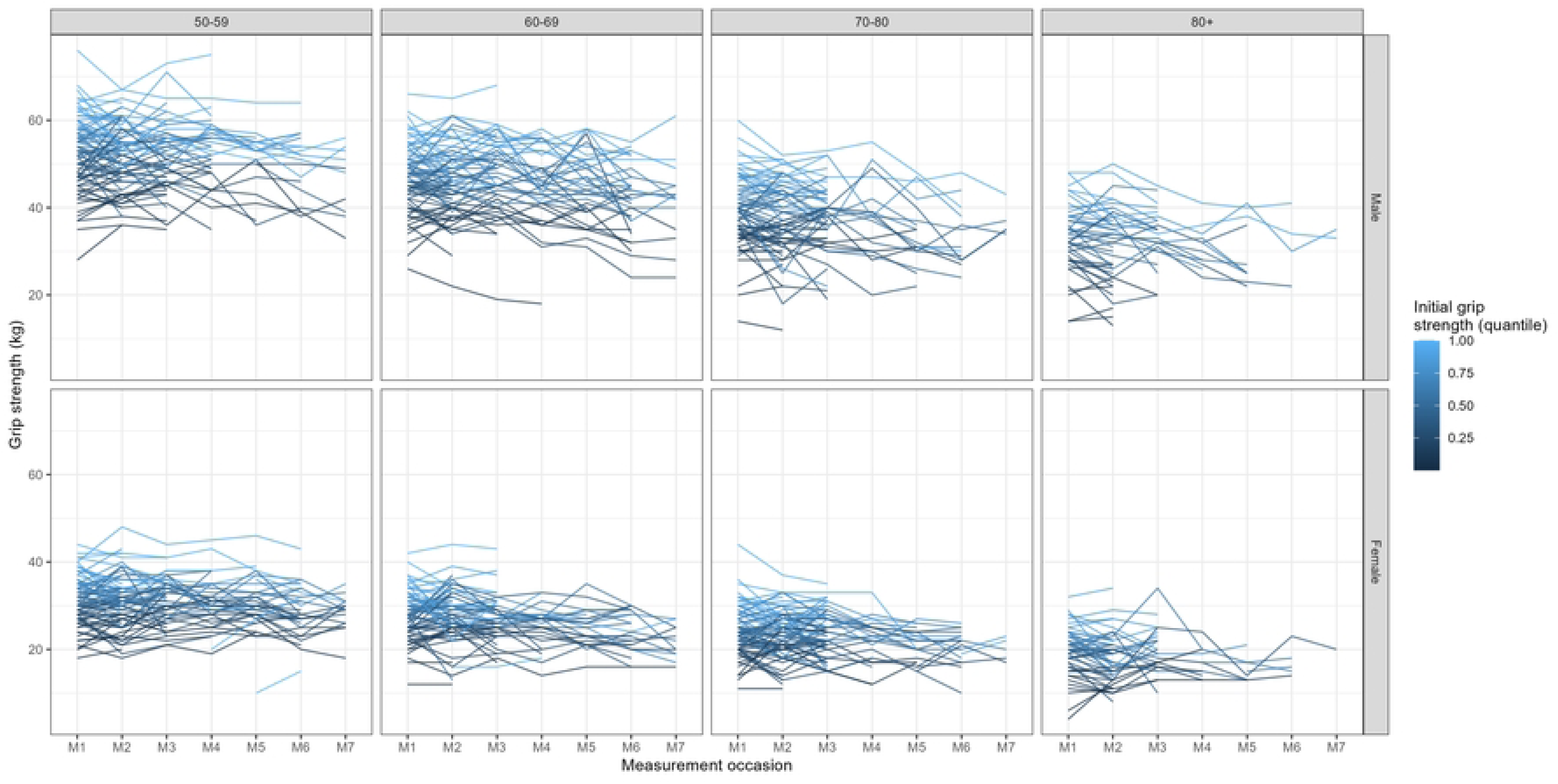
Profile plots of grip strength across measurement occasion, for a subset of participants. (the selection of 100 participants for each group is based on the quantile of grip strength at baseline).

**Fig 14.**
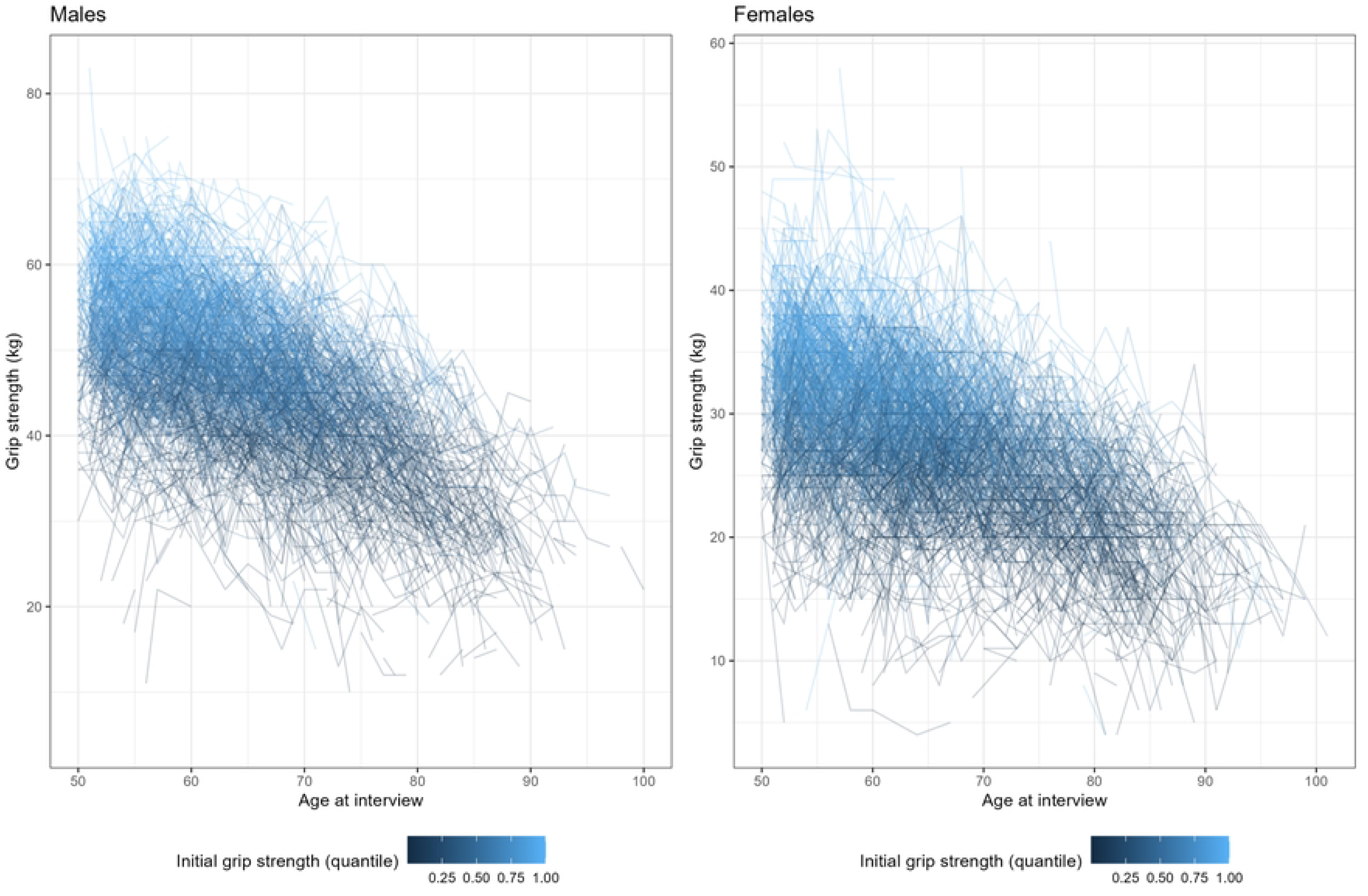
Profile plots of grip strength across measurement occasion for all participants, stratified by sex; age is used as time metric.

The profile plots highlight the trend towards the diminishing grip strength with age and show that the rate of change seems to accelerate over age (the slope at later ages is bigger than at the beginning). Older participants are followed up for shorter times, substantial increases or decreases in grip strength between measurement occasions can also be observed. The variability of the outcome tended to decrease at later measurement occasions, especially in the older age groups.

The scatterplots of the outcome measurements across waves and their correlations are shown using a generalized pairs plot (Fig. 11); across waves there were no substantial differences in the correlations (slightly lower in Wave 1) or variability of the outcome (Table 6).

The correlation of grip strength between measurements taken at different ages, indicated that the serial correlation was very high, generally above 0.70 for two-year periods and reduced slightly with the distance; correlations were generally slightly larger for males than for females (Fig. 15).

**Fig 15.**
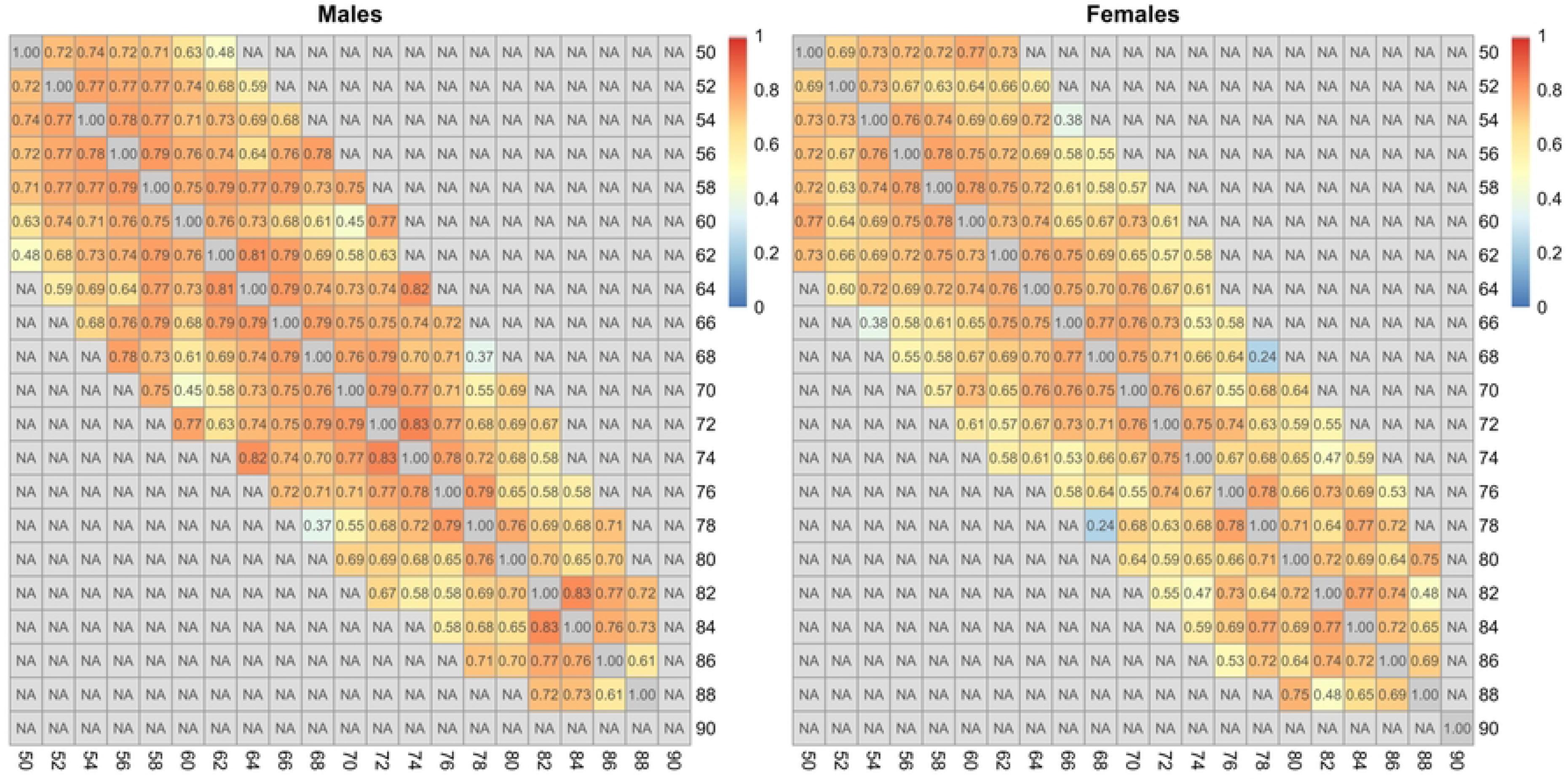
Correlation between successive outcome measurements taken at different ages in males and females; age of participants is grouped in two-year classes. Only estimates based on more than 20 observations are shown; the correlation between each pair of variables is computed using all complete pairs of observations on those variables.

Fig. 16 shows the smoothed estimated association between age and outcome for females, stratifying the data for grouped year of birth cohorts, and compares them to the estimates obtained using all longitudinal data, or cross-sectional data from only the first interview. Heterogeneity in the association between age and the outcome across year of birth cohorts were observed also for males, or considering different waves, and similar year of birth cohort effects could be observed for weight or height (online IDA report). These summaries should not be overintepreted, as they are not robust to missing data and assume independence between repeated measurements, but they suggest once again the potential importance of taking into account the year of birth cohort effect in the modelling, which can be addressed more formally during the modelling of the data.

**Fig 16.**
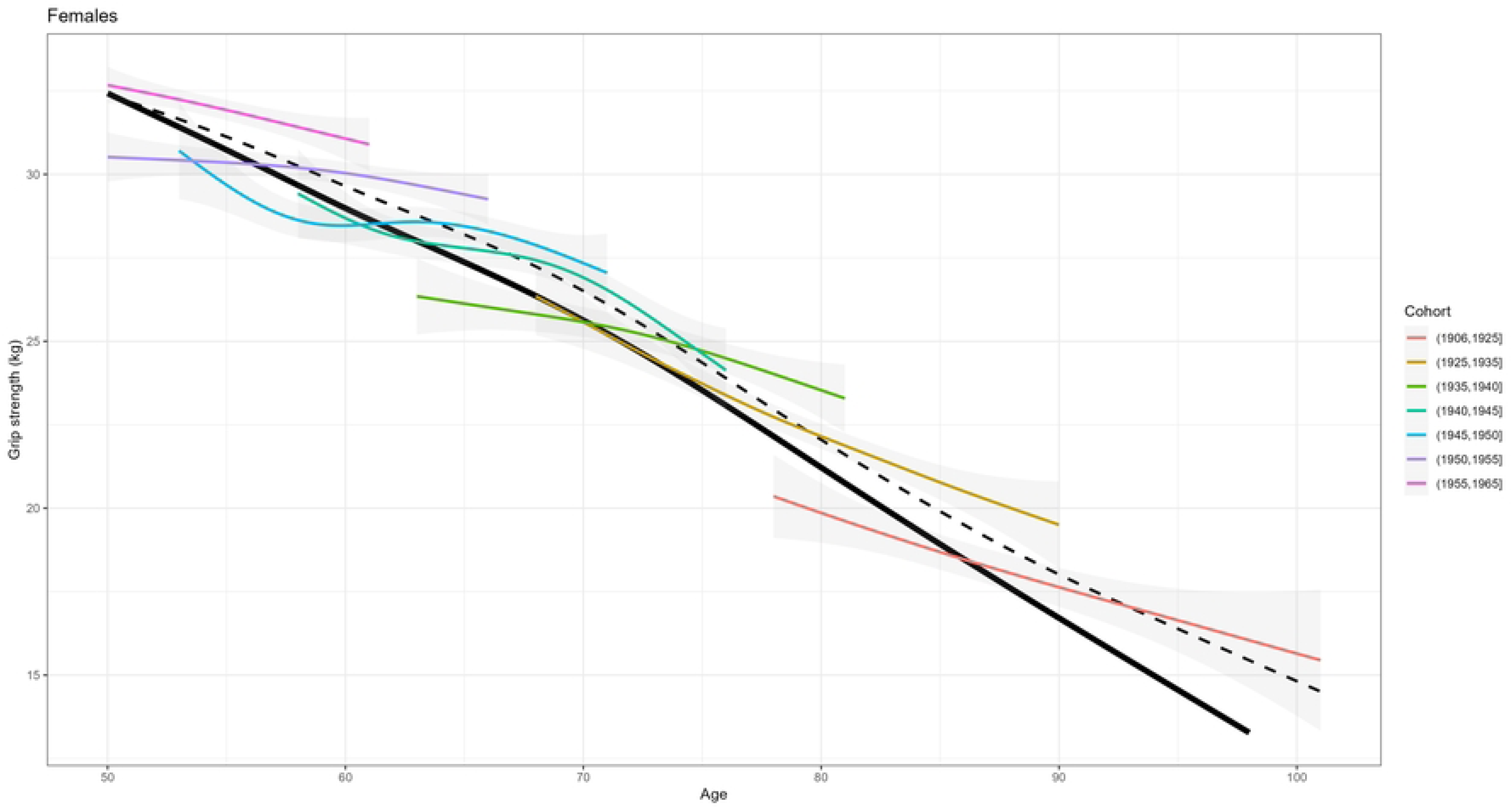
**Estimated association between age and grip strength within different subsets of data**; participants are stratified in grouped year of birth cohorts. Black lines are the estimates using all longitudinal data (dashed line) and cross-sectional data from the first interview (solid line).

Finally, the longitudinal changes of vigorous physical activity at least once a week is examined graphically, using a Sankey diagram (Fig. 17). The graph highlights that the transitions between active/not active state are common and that missing data are common (missing by design and losses to follow-up). These explorations are useful for providing domain experts a description of some of the characteristics of the sample that can be compared to the expected.

**Fig 17.**
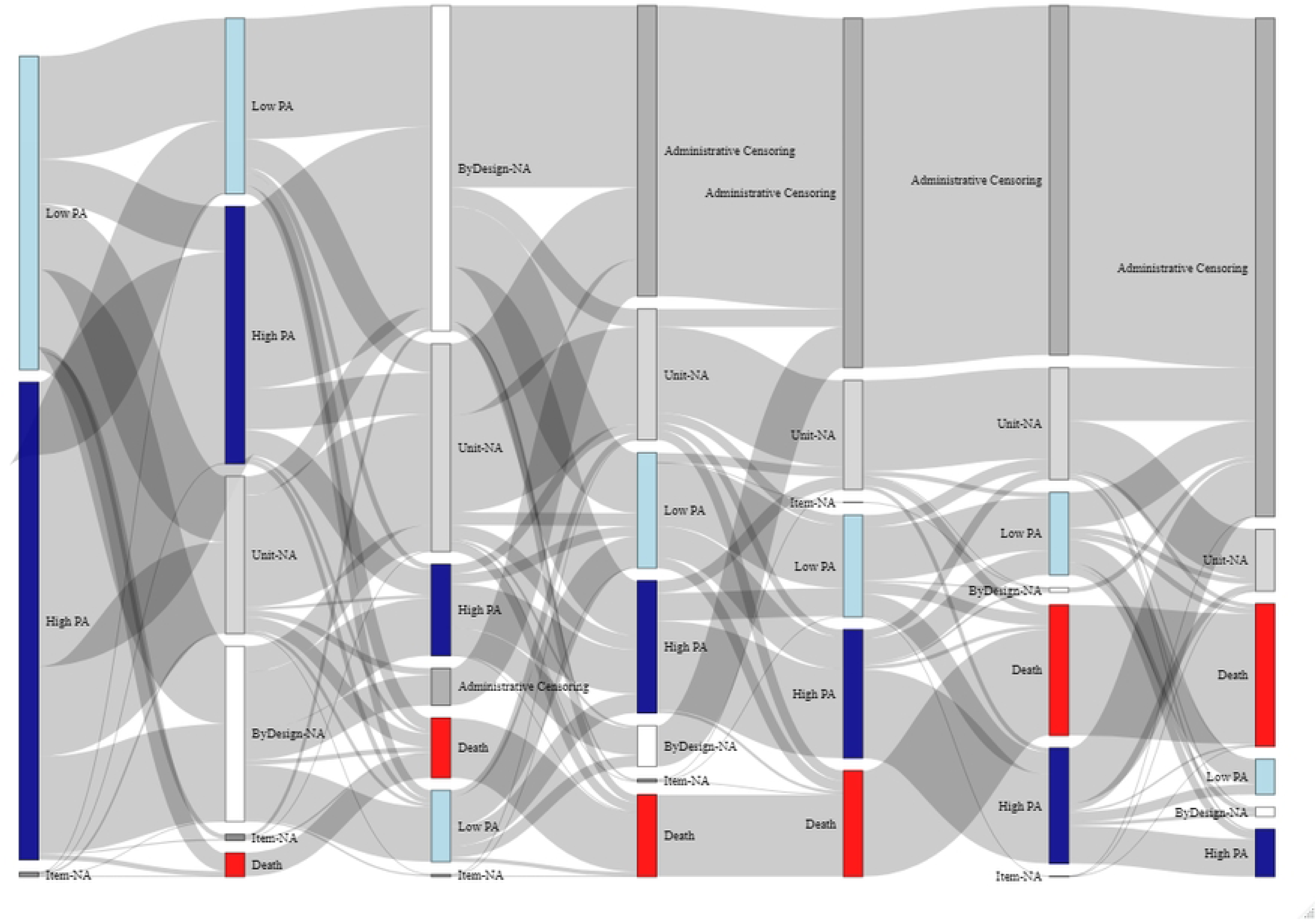
Sankey Diagram of vigorous activity status across measurement occasions. (all participants are displayed, with different reasons for missing values, measurement occasions are displayed from M1 (leftmost) to M7 (rightmost)).

### 4.5 Examples of potential consequences of data screening

Table 7 lists examples of how results from the IDA data screening could lead to new considerations for the data analyses.

**Table 7.** Potential consequences of data screening.

| Item | Topic | Consequences | Actions |
| --- | --- | --- | --- |
| **Participation profile** |  |  |  |
| P1 | Most participants had four or less measurement occasions (74%), 19% were measured only once. Therefore there was a lack of information for the identification of very flexible shapes of trajectories at the individual level. | Lack of information for the identification of very flexible shapes of trajectories at the individual level. | The number of random effects that can be included in the mixed model should be limited to three at most. The small number of repeated measurements may prevent from the inclusion of an autocorrelation process. |
| **Missing data** |  |  |  |
| M1<br>and<br>ME1 | Responders had substantially higher education than the target population, even when age and sex were taken into account. | If sampling bias is not taken into account, this could lead to lack of generalization to the entire population. | Statistical models need to account for the selection bias; this could be weighting approaches or adjustment for education. |
| M2 | About 20% of participants were lost to follow-up after first interview, about 35% after 12 years. Participants who dropped out of the study for reasons other than death had lower education and less healthy habits than those that remained in the study. | If the attrition mechanism is not appropriately taken into account in the statistical model, this could lead to biased results. | Methods that are robust to missing data mechanism are needed. With mixed models, the results will be robust to missing data predicted by the observations. Otherwise, joint models may be explored [15]. |
| ME2<br>and<br>ME3 | Deaths were common during the follow-up period in the study that includes an ageing population. For example, about 50% of the participants aged 80 or more at inclusion were dead after 6 years of follow-up. The trajectories of the outcome variable of participants that died differed from those that survived during follow-up. The characteristics of the participants that died were as expected, the quality of reporting of deaths was good. | If the deaths are not appropriately taken into, this could lead to biased results. | Random effect models can be used if deaths are assumed to be predictable by the observed outcome trajectories, while joint models with death as an event may assume a dependency based on unobserved outcomes values. Joint models for competing causes of dropout might be used, if both loss to follow-up and deaths are assumed to depend on the underlying outcome. A model assessing jointly the risk of dropout (possibly by nature of dropout - loss of follow-up or death) could be envisaged as a sensitivity analysis. |
| M4 | Missing values in the outcome among participants that were interviewed were not common, but the probability of their occurrence was larger for older participants and for females. | If missing values are not appropriately handled, this could lead to biased results. | This type of missing data (available interview, missing outcome) is handled as missing interviews in the statistical model. Mixed effect models, as mentioned above, will assume the missing data can be predicted from the observations. |

**Table 7.** Potential consequences of data screening
| Item | Topic | Consequences | Actions |
| --- | --- | --- | --- |
| M4 | Explanatory variables were missing by design in some waves/with some types of questionnaires. For example, current smoking was not available in the longitudinal questionnaire in later waves, physical activity variables were not measured in SHARELIFE interviews, and body mass was not measured in Wave 3. | Such missing values are not likely to introduce bias in the analysis, if handled by complete case analysis, as they are completely missing by design. However, the complete case analysis is not sensible, as the proportion of missing values is very large for some variables in some waves, which would result in decreased precision if incomplete cases are excluded from the analysis. | Imputation for missing values is needed for the explanatory variables missing by design. This finding may lead to possible changes in the analysis strategy, for example, using baseline values for smoking. |
| **Univariate descriptions** |  |  |  |
| U1,<br>U2 | There is a need for variable harmonization. The definition of some variables vary by wave/type of questionnaire (examples: current smoking, type of questionnaire used, questions that vary by wave, ...). | Introduction of errors (inconsistent definitions, avoidable missing values), measurement heterogeneity, reduction of statistical power, potentially information bias. | Data management to harmonize variables or adaptation of the statistical model to handle error of measurement and changes in measurement tool [16]. |
| U1 | Maximum grip strength was reported with terminal digit preference; digit preference were also observed for body mass and height. | Data include reporting errors that can lead to biased estimation and imprecision. | The impact of the errors on the results could be explored during modelling. Other studies showed that the SHARE data on grip strength is coarsened at random, and claimed that the consequences of rounding are minimal [17]. |
| **Multivariate descriptions** |  |  |  |
| V1 | Age was negatively associated to all other explanatory variables. | If the birth cohort is not taken into account, the estimated association between age and outcome might not represent the true association. | Inclusion of birth cohort/year of birth or year of first interview as explanatory variable in the statistical models. |
| **Longitudinal aspects** |  |  |  |
| L1<br>L2 | The rate of change in grip strength accelerated for older ages/older birth cohorts. The number of participants followed after 90 years of age was very limited. | Considering a statistical model linear in age misses the true functional form. Extrapolation of the results after 90 years old (especially in males) should be avoided. | Functional forms of age (e.g., quadratic, splines or fractional polynomials) should be investigated in statistical models for grip strength after adjusting for birth cohort. An indicator function for birth cohort or an interaction term for the age cohort with a function of time should be added as an explanatory variable. |

**Table 7.** Potential consequences of data screening
| Item | Topic | Consequences | Actions |
| --- | --- | --- | --- |
| L3 | The correlation of the outcome variable within each participant across measurement occasions was high, particularly for males. As expected, the correlations decreased for longer time periods between measurements. | If serial correlation is not taken into account, this could lead to biases estimation. | Carefully specify the correlation structure in the statistical model. For instance, considering only a random intercept in a mixed model would not be appropriate as it assumes that the correlation between the outcomes is constant and does not depend on the time lag between measurements. |
| L3 | The variance of the outcome decreases at older ages. | Decreasing variance over time can lead to biased estimation, if it is not taken into account. | The variance structure needs to be considered in a statistical model. For example, a random intercept alone in a mixed model would not be sufficient as it assumes that the variance of the outcome remains constant through time and random effects of the time functions should be considered. |
| L4 | Individuals change from high to low physical activity across measurement occasions or vice versa | The variation in the time-varying variable may affect the outcome. | The variation needs to be taken into account when interpreting the effect of physical activity and its association on the outcome grip strength. |

## 5 Discussion

IDA is crucial to ensure reliable knowledge about data properties and necessary context so that appropriate statistical analyses can be conducted and pitfalls avoided [3]. Often it is not transparent in publications what initial analyses researchers conducted and the reporting is poor. A multitude of decisions after examining data have an impact on results and conclusions [18].

An aim of IDA is to focus on the data properties that can justify the choice of statistical methods that rely on certain assumptions, and the findings provide additional indications for the interpretation and presentation of the results. For example, with longitudinal data when using mixed models, many different options can be used with random effects on different time functions and/or autocorrelated process; in a parametric model the IDA findings might suggest what basis of time functions would be the appropriate; the changes could be related to the choice of explanatory variables and the way in which they are modelled, to ways to account for informative dropouts or to adjust for covariates that may be associated to dropout and/or selection. Additionally, IDA could lead to the specification of sensitivity analyses.

For cross-sectional studies an IDA checklist for regression models with a continuous, count or binary outcome was developed by Heinze et al [4]. Several parts of such an IDA checklist carries over to longitudinal studies, for example as it relates to the univariate and multivariate descriptions of baseline characteristics, but there are additional IDA requirements that are specific to the longitudinal case, since measurements need to be examined at multiple time points and missing data need to be studied more thoroughly.

Some IDA elements are included in reporting guidelines such as the STROBE checklist [19]. These are items related to IDA data screening and items related to handling of consequences regarding expectations of the data. The IDA data screening elements that are included in the STROBE checklist comprise characteristics of study participants, number of missing participants, information about confounders, summary of follow-up time, and summary measures over time. Consequences are related to addressing potential sources of bias, methods for handling missing data, methods to control for confounding, methods to examine subgroups and interactions, sensitivity analyses.

It is important to remember that an IDA workflow is not a standalone procedure but is closely linked to the study protocol and the analysis strategy or the statistical analysis plan (SAP). A SAP describes the variables and outcomes that will be collected and includes ”detailed procedures for executing the statistical analysis of the primary and secondary variables and other data” [20]. Few elements of SAP are addressed in the STROBE reporting guidelines, which require reporting of any prespecified hypotheses, how the sample size was calculated, and the statistical methods used [21]. Guidelines for SAPs in clinical trials [22] and in observational studies [23] mention time points at which the outcomes are measured, timing of lost to follow-up, missing data, description of baseline characteristics and outcomes. By describing the statistical methods that the researcher chooses to use, the SAP addresses also issues related to how data properties will be explored and handled in the modeling (for example, describing how to handle missing data, correlated data, confouding, biases, sensitivity analyses, …). While researchers might anticipate the extent and patterns of missing data or potential sources of bias, a carefully conducted IDA workflow as proposed here can help researchers understand the data better and may also result in unanticipated findings. IDA can also identify errors and suggest better ways to conduct the analysis. It is suggested that changes to the analytic plan in response to IDA findings can be made, but should be properly documented and justified, to provide full transparency [24, 25].

Our IDA recommendations expand these guidelines and provide explanations and elaborations of the items that should be explored prior to undertaking the analysis detailed in the SAP. A fundamental principle of IDA is to explicitly avoid hypotheses generation activities [1]. Since IDA findings could lead to changes in the analysis strategy, a SAP needs to include both an IDA plan and details of the analysis strategy for transparency and reproducibility and to avoid ad-hoc decisions. Thus, associations of explanatory variables defined in the analysis strategy with the outcome variable are excluded from the IDA plan. However, in longitudinal studies, IDA involves the description of trends; summarizing the outcome variable across time or visualizing profiles are indispensable for these types of studies.

In longitudinal studies the time metric has to be clearly defined. Time since enrollment, calendar time, or measurement occasions are commonly used time metrics. Participation needs to be carefully examined, namely who the participants are, when they enter the study, when and why they leave, as well as drop-out effects. This has consequences for statistical analyses choices about handling missing data, random effects, or competing risks. Missing data are a major challenge in longitudinal studies and their exploration provides a deeper understanding of the data. It is expected that a SAP specifies how missing values will be handled in the analyses [26] but choices might depend on the missing data characteristics that could be revealed in an IDA report and could provide insights about assumptions and approaches for handling missing data. Variables need to be examined at multiple time points; profile plots for time-varying variables can be helpful to summarize changes and variability of variables within participants as well as possible longitudinal trends across participants. Longitudinal trends, correlations and variability, and period or cohort effects may also be examined.

In the longitudinal setting IDA explorations can quickly become overwhelming even with a small numbers of variables. Our IDA checklist may be useful to guide researchers to carefully consider topics. A worked example with available data and reproducible R code including many effective data visualizations is provided, which can be adapted for developing IDA plans for other studies. We believe that the checklist, the worked example and the R code will help data analysts in planning and performing IDA data screening for longitudinal studies to provide researchers with a better understanding of their dataset.

To summarize, we provide recommendations for a check list for IDA data screening in longitudinal studies with examples for data visualizations to enable researchers follow a systematic approach and reproducible strategies. Such an IDA improves the understanding of opportunities and shortcomings of a dataset, addresses the problem of model assumptions and enhances the interpretation of model results.

## Supporting information

**Supplementary file 1 IDA plan for the application presented in the paper.**

Detailed description of the IDA plan.

**Supplementary file 2 Additional tables and figures.**

## Data Availability

Data are available for research purposes at https://share-eric.eu/data/data-access

https://stratosida.github.io/longitudinal/

## Acknowledgments

This work was developed as part of the international initiative of Strengthening Analytical Thinking for Observational Studies (STRATOS). The objective of STRATOS is to provide accessible and accurate guidance in the design and analysis of observational studies (http://stratos-initiative.org/). Members of the Topic Group Initial Data Analysis of the STRATOS Initiative are Mark Baillie (Switzerland), Marianne Huebner (USA), Saskia le Cessie (Netherlands), Lara Lusa (Slovenia), Carsten Oliver Schmidt (Germany).

L.L. was partially supported by ARRS research program P3-0154.

